# Automated and interoperable methods for generalizable development of clinical machine-learning models for predicting neuromorbidity in critically ill children

**DOI:** 10.1101/2025.08.01.25332805

**Authors:** Ruoting Li, Christopher M. Horvat, Mehdi Nourelahi, Eddie Pérez Claudio, Jason Hammett, Mark S. Wainwright, Robert S.B. Clark, Alicia K. Au, Harry Hochheiser

**Affiliations:** Department of Critical Care Medicine, University of Pittsburgh, Pittsburgh, Pennsylvania, USA; Safar Center for Rehabilitation Research, University of Pittsburgh, Pittsburgh, Pennsylvania, USA; Department of Biomedical Informatics, University of Pittsburgh, Pittsburgh, Pennsylvania, USA; AWS, Dublin, Ohio, USA; Division of Pediatric Neurology, Seattle Children’s Hospital, Seattle, Washington, USA

**Keywords:** Pediatric Neurological morbidity, Clinical Decision Support, FHIR, Cloud-Based Analytics, Predictive Modeling

## Abstract

**Objectives:** To streamline the development of clinical machine learning (ML) models for predicting acute neurological morbidity in critically ill children by extending our prior work to create a standardized, reproducible, and scalable workflow leveraging Fast Healthcare Interoperability Resources (FHIR), cloud infrastructure, and automated ML tools.

**Methods:** We developed workflow for extracting, cleaning, and modeling pediatric intensive care unit (PICU) data, using 168 biomarkers from 7,403 encounters at an academic Children’s hospital between 2020 and 2024. Data were processed and stored in a compliant, secure cloud environment. We evaluated four feature sets: a baseline set from prior work, a complete set, a filtered set, and a light gradient boosting machine (LightGBM)-selected set for prediction of acquired neurological morbidity. Automated ML was used to train, validate, and deploy models, with performance assessed using the area under the receiver operating characteristics curve (AUROC), area under the precision recall curve (AUPRC), F1 score, calibration metrics, and Shapley additive (SHAP) values. A FHIR-based version of the pipeline was also implemented and evaluated on a 2020 subset of the cohort.

**Results:** Filtered and LightGBM-based feature sets achieved the highest predictive performance, with AUROCs of 0.90 (95% CI: [0.88-0.92]) for both, and AUPRCs of 0.68 (95% CI: [0.63-0.73]) and 0.67(95% CI: [0.62-0.72]), respectively. SHAP value analysis revealed consistent top features across models, with vital signs and key laboratory values prominently ranked. Models trained using FHIR-formatted data from a 2020 cohort (n = 1,339) demonstrated comparable performance to those built on the complete dataset, with an AUROC of 0.87 (95% CI: [0.81-0.93]).

**Conclusions:** This study demonstrates the feasibility of a cloud-compatible, standards-based approach to clinical ML model development. By leveraging interoperable data formats and automated modeling workflows, this approach supports scalable, reproducible model construction and evaluation, enabling improved efficiency and transparency in clinical decision support.

## 1. Background and Significance

New approaches are needed to standardize and formalize the development of machine learning algorithms for clinical decision support. Our recent work on developing interoperable machine learning models to predict neurologic morbidity in critically ill pediatric patients demonstrates both the promise and complexity inherent in clinical predictive modeling.^1^ Despite achieving robust external validation and biomolecular corroboration, the model development process involved extensive data extraction from heterogeneous clinical data warehouses, variable harmonization, feature engineering, and complex workflows implemented in diverse computational environments. This experience is emblematic of common experience in this area; developers of these tools face the challenges of extracting data from clinical data warehouses, harmonizing data from different sources, using machine learning tools and, and cleaning data, often while working in restricted computational environments as needed to protect patient privacy. Even when data is provided in standard data formats such as the OMOP common data model,^2^ data cleaning, imputation, and modeling steps often rely on customized workflows. As a result, each new model or new environment requires significant customization. To realize the benefits of machine learning for clinical decision support, new approaches are needed to minimize repeated effort, increase opportunities for code reuse and reproducibility, support frameworks capable of informing ongoing evaluation and evolution of models, and facilitate integration with operational systems used by health care systems.^3–5^ We propose replacing bespoke components and workflows with an end-to-end, interoperable pipeline to accelerate and improve the rigor of model development and deployment.

Advances in data and computing tools provide possibilities for improved processes. Fast Health Interoperability Resources (FHIR) data stores provide data in a predictable and consistent format, particularly when complaint with the federally mandated United States Core Data for Interoperability (USCDI) specifications.^6^ Emerging bulk FHIR capabilities in electronic health record (EHR) systems support the extraction of the large amounts of data necessary to train and validate models.^7^ Software evolution has led to the development of powerful toolkits capable of enabling model development at a higher-level of abstraction. Just as tools like scikit-learn,^8^ Tensor Flow,^9^ and Pytorch^10^ simplified the construction of models, next-generation automated machine learning (AutoML) tools manage the details of potentially complex steps such as data imputation and hyperparameter tuning.^11^

## 2. Objectives

To build on our work in developing models for predicting adverse acquired neurological morbidities in pediatric intensive care unit (PICU) patients,^1^ by constructing a workflow establishing an end-to-end process for building a data model, including data access from a FHIR store, data cleaning, modeling, testing, validation, reporting, and provision of an inference endpoint, in preparation for a prospective, multicenter clinical validation study. This workflow provides a reusable and adaptable framework capable of supporting ongoing quality assurance, as well as generalizing to other predictive targets.

## 3. Methods

Our original model used 45 structured EHR variables including vital signs, lab test results, medications, and patient age, to predict risk of neurological morbidity defined as a composite outcome including orders for brain function tests (electroencephalogram [EEG], magnetic resonance imaging [MRI], or head computerized tomography [CT]-scan) or indicators of delirium. Up to 48 hours of data for each patient were used, with a 12-hour censored time horizon before any event. Temporal features were converted into summaries using trend indicators (min, max, average, etc.) prior to feature selection via information gain. The best-performing XGBoost model had an area under the receiver operating characteristic curve (AUROC) of 0.89.^1^

Our revised model extends this effort, through a more systematic approach to feature-selection, implementation as a FHIR-based service, and algorithmic enhancements including the use of an auto-ML strategy for optimizing model development.

This study was approved by the University of Pittsburgh’s Human Research Protection Office (STUDY20050220). No registered protocol was prepared for this study. There was no patient or public involvement in the design of this study. As clinical data is involved, data can be shared only with a data use agreement with the University of Pittsburgh. Source code is available upon request.

### 3.1. Data Acquisition and Preparation

Data for PICU visits from UPMC Children’s Hospital occurring between January 1, 2020 and November 8, 2024 were extracted from an internal data warehouse and stored in a health insurance portability and accountability act (HIPAA)-compliant secure cloud environment. Records for patients without a documented age, or with an age of greater than 300 months were excluded. Extracted data included laboratory tests, medications, demographics, and vital signs, as detailed in our previous work.^1^ We expanded on our earlier work, using a more systematic approach to identify possible features. We began with all available observation resources, including 1,487 unique LOINC codes. A semi-automated curation pipeline was then applied to select relevant features. First, a rule-based filter retained codes with more than 10,000 observations. Next, a human-in-the-loop method was used to classify data types: codes with non-empty values that could be fully cast to numeric format were automatically classified as numeric; those with fewer than 30 unique values were classified as categorical. The remaining codes were manually reviewed by clinical experts to determine appropriate data types. For each LOINC code, summary statistics reports were generated. Clinical experts used these reports to identify variables to retain, specify data cleaning steps for unusual formats, and define curation rules for boundary handling. Orders for head CT, brain MRI, or EEG, or indicators of treated delirium (behavioral health consult and/or administration of dexmedetomidine, olanzapine, haloperidol) were used to define clinical suspicion of neurological morbidity. Other medications and LOINC codes were used as predictive features when possible.

The resulting dataset was cleaned by converting unusual formats such as text strings or mathematical qualifiers to computable values, removal or truncation of outliers, and grouping of relevant variables such as pupillary reaction and ventilator make/model to clinically meaningfully categories. Thresholds used for outlier removal and truncation in numerical variables are detailed in **Supplemental Table 1**. All records were deduplicated.

An observation window was defined for each eligible encounter. For encounters with neurological morbidities, the earliest observation time of any of the components of the definition of neurological morbidity was used as the reference event time. To enable sufficiently early detection, the observation window was defined as ending 24 hours before that event. Data from up to 48 hours before that point was included in the window. Encounters were excluded if the time from arrival to the event was less than 24 hours, or if no relevant observations were available within the defined observation window. Encounters without clinical suspicion for adverse neurological morbidity were excluded if the length of stay was less than 48 hours. For those that remained, a random 48-hour observation window was selected. **Figure 1** illustrates how observation windows were determined for each encounter type.

**Figure 1.**
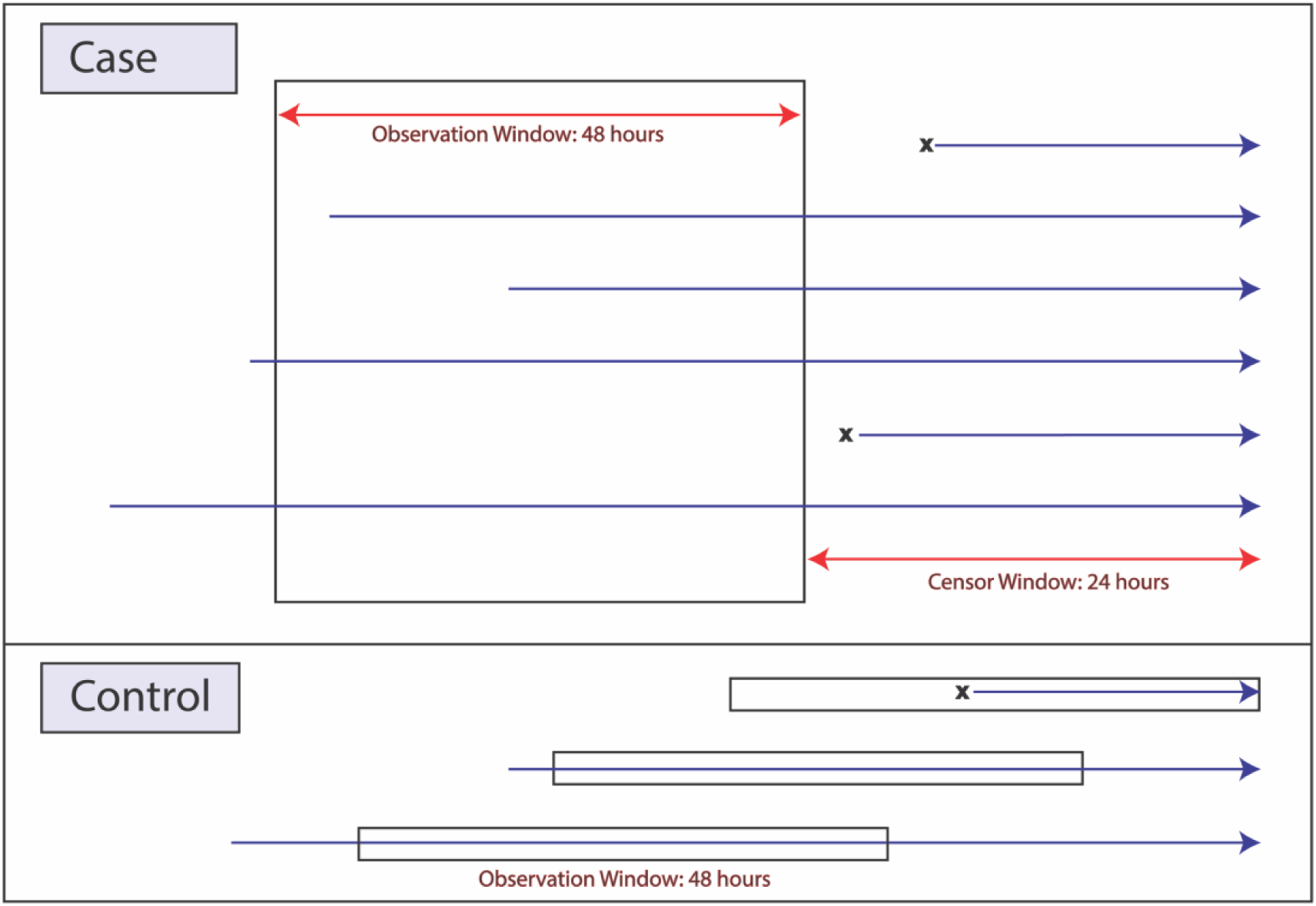
Illustration of observation and censor window definitions for encounters with and without neurological morbidity. For cases (top), the 48-hour observation window ends 24 hours before the earliest outcome indicator (censor window). For controls (bottom), a random 48-hour window is selected during the hospital stay. Encounters marked with an “X” indicate exclusion due to stay duration or insufficient length of observation.

Cases were divided into temporally defined training and test datasets, with cases between January 1, 2020 and December 31, 2022 forming the training set and cases from January 1, 2023 to November 8, 2024 forming the test dataset.

### 3.2. Feature Extraction

Each numeric time-series indicator was converted into 19 non-temporal summary statistics (first. value, last value, slope, median, mean, etc.).^12^ Categorical features were discretized into four values (first, last, second-to-last, and mode). Binary biomarkers such as medication or ventilation were coded as 1 if present at any time during the observation window, 0 otherwise. Categorical biomarkers with no readings recorded during the 48-hours window were coded with a distinct “missing” indicator.

We performed a series of experiments aimed at exploring the utility of including features beyond those used in our original published effort. We explored four sets of features. A baseline feature set contained the features used in our previously published work.^1^ An agnostic feature set used all available features. A filtered feature set narrowed the set of features from the complete set, removing all biomarkers with more than 80% missing values and weak (<0.2) correlation with elements of the composite definition of neurological morbidity. The filtered dataset also involved simplification of character of cough and body position, the two categorical biomarkers with more than 20 values. For these biomarkers, the 20 most frequently found values were retained and all remaining entries were summarized with the value “other”. Finally, the LightGBM feature set used a LightGBM^13^ feature-selection process provided by Amazon Web Services (AWS) AutoML^14^ to select features.

### 3.3. Models

Automated ML platforms provide end-to-end tools for constructing machine learning models, wrapping steps including feature selection, algorithm selection, model construction, internal validation, hyperparameter tuning, and evaluation into a single workflow.^11^ Available in both numerous open-source packages^15^ and through major cloud service providers, AutoML tools are a further step toward streamlining the process of building ML models, eliminating the need for rewriting of code for train, test, and validation and providing greater reproducibility.

We used the AutoML functionality provided by AWS^14^ to build models, using five-fold cross-validation to evaluate each model, storing the best performing pipeline in an Amazon S3 storage bucket, and registering the resulting model in the Amazon SageMaker Model registry.^16^ This model was then deployed to an inference endpoint supporting real-time prediction. This endpoint was used to evaluate performance with a hold-out dataset.

We evaluated six key metrics: accuracy, precision (positive predictive value or PPV), recall (sensitivity), F1 score, AUROC, and area under the precision-re all curve (AUPRC) on the training, cross-validation, and test sets. SHAPley values^17^ were used to identify biomarkers exerting the largest influence on predicted neurological morbidities. Brier scores were used to assess model calibration.

### 3.4. Fast Health Internet Resources Conversion and Modeling

To demonstrate the potential utility of FHIR as the basis for building and providing machine learning clinical decision support models, we developed a FHIR version of our predictive models. We created a Python script to translate our data from tabular from into four core FHIR resources: Patient, Encounter, Observation, and MedicationAdministration.^18^ We stored the resulting FHIR resources in AWS HealthLake,^19^ a HIPAA-compliant FHIR service. We then used HealthLake query services to extract data for use in our modeling pipeline. Using a subset of the data from 2020, we recreated the four experiments described above.

## 4. Results

### 4.1. Data Acquisition and Preparation

Our initial cohort included 12,110 encounters. After exclusion due to age and data availability constraints, the final cohort included 7,403 encounters. Details of the cohort selection are given in **Supplemental Figure 1**. Demographics comparison of the case and control groups for the entire dataset and for the training (1/1/2020-12/31/2022) and test datasets (1/1/2023-11/8/2024) are given in **Table 1**. In all cases, groups were comparable in age, patient gender, and Glasgow coma score, a measure of consciousness used in intensive care units.^1^ Cases were significantly higher in mechanical ventilation status, presence of endotracheal tube, use of vasoactive medication, and use of sedative-analgesic medication.

**Table 1.** Demographic characteristics of the case and control encounters across entire cohort and the training and test datasets. *Significant difference between case and control (p <0.05) ^1^All patients were identified as either male or female ^2^The last recorded Glasgow coma scale score for the encounter prior to the censored time horizon ^3^Vasoactive medications include dobutamine, dopamine, epinephrine, norepinephrine, or milrinone ^4^ Sedative-analgesic medications include fentanyl, hydromorphone, midazolam, or morphine Abbreviations: IQR, interquartile range Notes: Continuous variables were compared using Welch’s t-test; categorical variables were compared using Chi-square tests.

|  | Entire Cohort |  | Train |  | Test |  |
| --- | --- | --- | --- | --- | --- | --- |
|  | Case | Control | Case | Control | Case | Control |
| **Size** | n=887 | n=6,516 | n=577 | n=4,205 | n=310 | n=2,311 |
| **Age (months), median (IQR)** | 74 (15,177) | 78 (20,166) | 74 (14,174) | 75 (18,165) | 73 (16,186) | 83 (24,168) |
| **Female, n (%)<sup>1</sup>** | 405 (46) | 3,058 (47) | 264 (46) | 1,980 (47) | 141 (45) | 1,078 (47) |
| **Glasgow Coma Scale Score, median (IQR)<sup>2</sup>** | 15 (10,15) | 15(14,15)* | 15(10,15) | 15(14,15)* | 14(10,15) | 15(14,15)* |
| **Mechanical Ventilation, n (%)** | 329(37) | 1,414(22)* | 215(37) | 876(21)* | 114(37) | 538(23)* |
| **Endotracheal Tube, n(%)** | 251(28) | 378(6)* | 164(28) | 234(6)* | 87(28) | 144(6)* |
| **Vasoactive Medication, n(%)<sup>3</sup>** | 132(15) | 292(5)* | 86(15) | 200(5)* | 46(15) | 92(4)* |
| **Sedative-Analgesic Medication, n (%)<sup>4</sup>** | 410(46) | 1,012(16)* | 267(46) | 657(16)* | 143(46) | 355(15)* |

### 4.2. Feature Extraction

Our four feature sets raised in size from the 44 biomarkers and 584 summarized features used in our original model^1^ to the 168 biomarkers and 2,204 summarized features in the complete set (**Table 2**). One binary biomarker, Emotional Support Management, was coded as a categorical biomarker for the complete feature set only. The biomarkers represented in the filtered feature set included all the features in the baseline data set and most (84/91) of those included in the LightGBM feature set. Of the 44 biomarkers included in the baseline feature set, 22 were included in the LightGBM feature set (**Supplemental Figure 2, Supplemental Figure 3; a complete list of features in each of the four sets is provided in Supplemental data**).

**Table 2.** Distributions of biomarkers and features in the four feature sets.

| Feature Set | Biomarkers | Features |
| --- | --- | --- |
| Baseline | 44 | 584 |
| Complete | 168 | 2,204 |
| Filtered | 128 | 1,659 |
| LightGBM | 91 | 525 |

### 4.3. Models

**Table 3** summarizes the predictive performance of the AutoML Models over the four experiments. In all cases, the XGBoost models were selected by the AutoML as the best performing models. The models performed strongly during training, particularly for the filtered and LightGBM feature sets. The validation and test performance remained consistent, especially in terms of accuracy and AUROC. Using the default decision threshold of 0.5, the filtered feature set achieved the highest recall of 0.64 (95% CI: [0.58-0.69]) and F1 score of 0.63 (95% CI: [0.59–0.68]). This result is expected, as this set utilized 1,659 features from 128 biomarkers selected through filtering, which likely offered a balance between reduced noise (compared to the complete feature set) and richer feature representation (compared to the smaller baseline and LightGBM feature sets).

**Table 3.** AutoML model performance across training, validation, and test datasets for the four experimental settings. Performance metrics are reported as value (95% confidence interval) for the training and test sets only. Validation performance is reported by AutoML using cross-validation and does not include AUPRC. Performance on the training and test sets was evaluated via the inference endpoint. The best results on the test set for each metric are bolded.

| Feature Set | Baseline |  |  | Complete |  |  | Filtered |  |  | LightGBM |  |  |
| --- | --- | --- | --- | --- | --- | --- | --- | --- | --- | --- | --- | --- |
|  | Train | Validation | Test | Train | Validation | Test | Train | Validation | Test | Train | Validation | Test |
| **Accuracy** | 0.96<br>(0.96-0.97) | 0.89 | 0.90<br>(0.89-0.91) | 0.98<br>(0.98-0.99) | 0.92 | 0.90<br>(0.88-0.90) | 1.00<br>(1.00-1.00) | 0.92 | 0.91<br>(0.90-0.92) | 0.99<br>(0.99-1.00) | 0.920 | **0.90**<br>**(0.89-0.91)** |
| **Precision** | 0.78<br>(0.74-0.80) | 0.53 | 0.57<br>(0.52-0.62) | 0.88<br>(0.86-0.91) | 0.65 | 0.52<br>(0.48-0.57) | 0.98<br>(0.97-0.99) | 0.70 | **0.63**<br>**(0.58-0.69)** | 0.95<br>(0.93-0.97) | 0.685 | 0.58<br>(0.53-0.63) |
| **Recall** | 0.97<br>(0.95-0.98) | 0.63 | 0.65<br>(0.60-0.70) | 0.98<br>(0.97-0.99) | 0.65 | **0.65**<br>**(0.60-0.70)** | 1.00<br>(1.00-1.00) | 0.62 | 0.64<br>(0.58-0.69) | 1.00<br>(1.00-1.00) | 0.626 | 0.62<br>(0.57-0.67) |
| **F1 Score** | 0.86<br>(0.84-0.88) | 0.58 | 0.61<br>(0.56-0.65) | 0.93<br>(0.91-0.94) | 0.65 | 0.58<br>(0.54-0.62) | 0.99<br>(0.99-1.00) | 0.66 | **0.63**<br>**(0.59-0.68)** | 0.98<br>(0.97-0.98) | 0.653 | 0.60<br>(0.56-0.64) |
| **AUROC** | 0.99<br>(0.99-0.99) | 0.87 | 0.88<br>(0.86-0.90) | 1.00<br>(1.00-1.00) | 0.91 | **0.90**<br>**(0.88-0.92)** | 1.00<br>(1.00-1.00) | 0.90 | 0.90<br>(0.88-0.92) | 1.00<br>(1.00-1.00) | 0.905 | 0.90<br>(0.88-0.92) |
| **AUPRC** | 0.94<br>(0.93-0.96) |  | 0.66<br>(0.61-0.71) | 0.99<br>(0.98-0.99) |  | 0.66<br>(0.61-0.72) | 1.00<br>(1.00-1.00) |  | **0.68**<br>**(0.63-0.73)** | 1.00<br>(0.99-1.00) |  | 0.67<br>(0.62-0.72) |

The four experiments yielded AUROC values of 0.88 (95% CI: [0.86–0.90]) for Baseline, 0.90 (95% CI: [0.88–0.92]) for Complete, 0.90 (95% CI: [0.88–0.92]) for Filtered, and 0.90 (95% CI: [0.88–0.92]) for LightGBM. The corresponding AUPRC values were 0.66 (95% CI: [0.61–0.71]), 0.66 (95% CI: [0.61–0.72]), 0.68 (95% CI: [0.63–0.73]), and 0.67 (95% CI: [0.62–0.72]), respectively. The complete feature set achieved the highest AUROC, while the filtered set achieved the highest AUPRC. Additionally, the LightGBM feature set slightly outperformed the baseline feature set in both metrics. Tradeoffs between PPV, Sensitivity, and F1 score across the four scenarios are illustrated in **Supplemental Figure 4**, AUROC and AUPRC results can be found in **Supplemental Figures 5 and 6**. Hyperparameters selected by the AutoML tuning are found in Supplemental Table 2.

Models were generally well-calibrated, with Brier scores for the four feature sets of 0.078, 0.080, 0.067, and 0.072, for the baseline, complete, filtered, and LightGBM feature sets, respectively, aligning with their overall predictive performance. The calibration curves fall below the diagonal line, particularly in the mid-range of predicted probabilities (0.4–0.8), indicating that the AutoML models tend to be overconfident in this range. Calibration details are show in **Supplemental Figure 7**.

### 4.4. Shapley values and missingness

Models built using the four feature sets showed significant consistency in the lists of features with the highest Shapley values. Of the 15 highest-rank features for each of the four feature sets, only six were found in only one of the four models, and three of those were in the baseline feature set which started from a significantly smaller set of biomarkers. Blood pressure (mean blood pressure [MBP], diastolic blood pressure[DBP], systolic blood pressure [SBP]), Sp02, Pulse, Temperature, Respiratory Rate, and Blood urea nitrogen were highly ranked for all four feature sets. Six additional biomarkers (Platelets, Peds Coma Score, Phosphate (blood/serum), Inhaled oxygen flow rate, and Humpty Dumpty Score) were found in three of the four lists of the 15 highest-ranked features (**Figure 2**). Missingness for top-ranked features was low. Of the 22 features identified as among the top 15 Shapley values for at least one of the four feature sets, only four (INR, 61.4% missing; Inhaled oxygen flow rate, 31.2%; Platelets, 14.08%; and White blood cell count, 13.9%) were missing more than 10% of the time. Of these four, INR and White blood cell count were among the top 15 only for the baseline dataset (**Table 4**). Full details of missingness rates for all features are given in **Supplemental tables 3, 4, and 5. Supplemental Table 6** groups biomarkers insight into which seven broad categories, and reports the average missingness within each category and the categorization of the top biomarkers.

**Figure 2.**
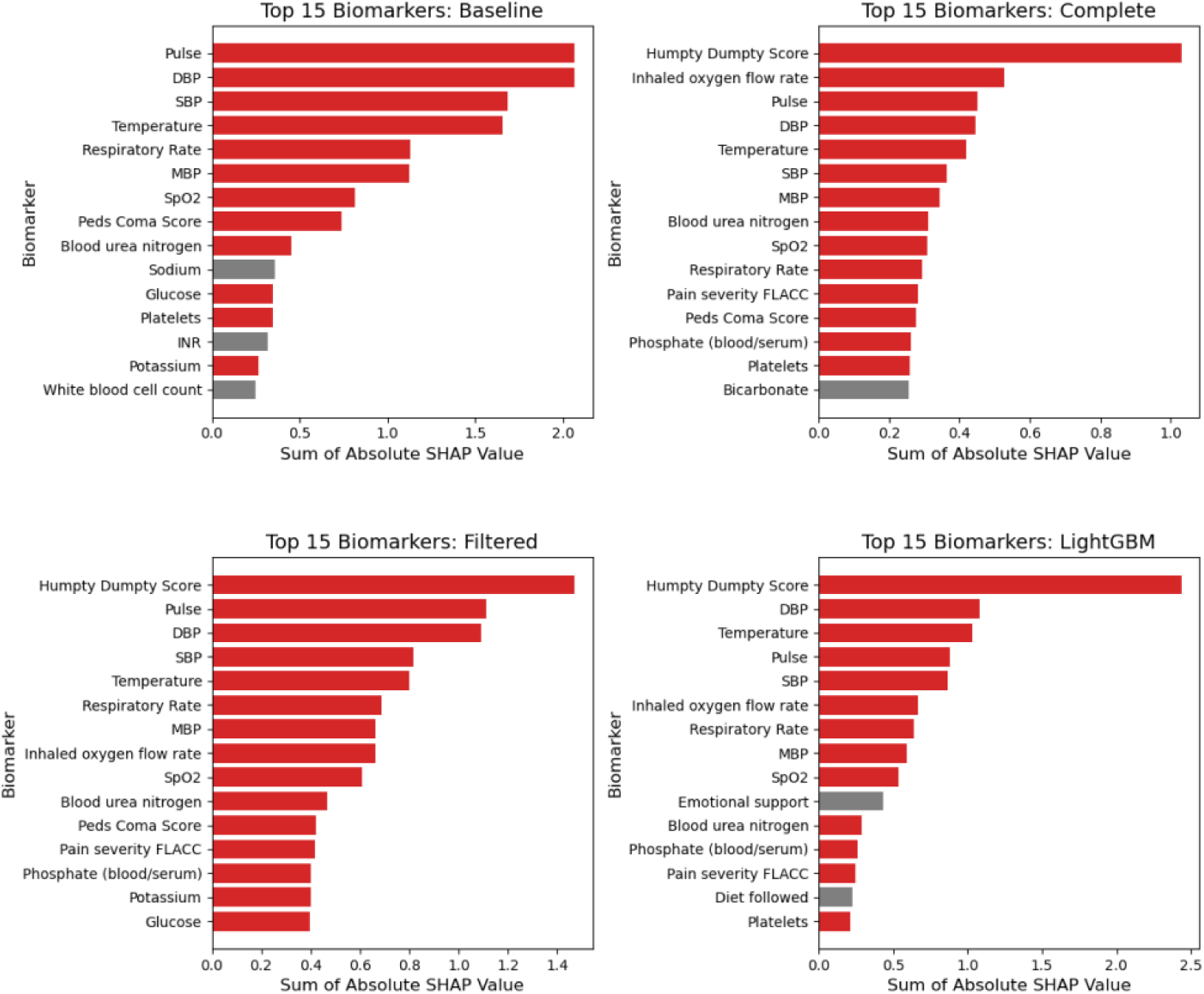
Shapley values for the top 15 features in each feature set, ranked by descending magnitude of Shapley value. Features occurring in more than one feature set are drawn in red. *Abbreviations*: DBP, diastolic blood pressure; FLACC, face, legs, activity, cry, consolability scale; INR, international normalized ratio; MBP, mean blood pressure; SBP, systolic blood pressure; SpO_2_, oxygen saturation.

**Table 4.** Missingness rates for the 22 biomarkers included in the union of the top (n=15) biomarkers with the highest Shapley values for each of the four feature sets (N = 7,403 encounters). For each biomarker, the number and % of encounters without values is reported.

| Biomarker_Name | Type | # Without values | % without values |
| --- | --- | --- | --- |
| Pulse | Numeric | 11 | 0.15 |
| DBP |  | 11 | 0.15 |
| SBP |  | 11 | 0.15 |
| Temperature |  | 11 | 0.15 |
| Respiratory Rate |  | 11 | 0.15 |
| MBP |  | 13 | 0.18 |
| SpO2 |  | 11 | 0.15 |
| Peds Coma Score |  | 1 | 0.01 |
| Blood urea nitrogen |  | 254 | 3.43 |
| Sodium |  | 197 | 2.66 |
| Glucose |  | 234 | 3.16 |
| Platelets |  | 1042 | 14.08 |
| INR |  | 4549 | 61.45 |
| Potassium |  | 250 | 3.38 |
| White blood cell count |  | 1030 | 13.91 |
| Humpty Dumpty fall risk assessment score |  | 203 | 2.7 |
| Inhaled oxygen flow rate |  | 2309 | 31.2 |
| Pain severity total [Score] FLACC |  | 639 | 8.6 |
| Phosphatein Serum or Plasma |  | 474 | 6.4 |
| Bicarbonate |  | 205 | 2.77 |
| Emotional support management | Binary | 70 | 1.0 |
| Diet followed | Categorical | 11 | 0.2 |

### 4.5. Fast Health Internet Resources Conversion and Modeling

We replicated our modeling pipeline using FHIR data from a store of 1,339 encounters from 2020. Encounters occurring from January 1 – September 30 (n=1, 026) were used to train the models, with remaining encounters (n=313) from October 1 – December 31 used as a hold-out test set (**Table 5**). Due to the smaller sample size, there is a slightly larger discrepancy between validation and test performance for threshold-dependent metrics such as F1 score (**Table 6**). However, accuracy and AUROC remained consistent across datasets, with results comparable to those observed in models developed using the full cohort. LightGBM showed the best performance, with a recall of 0.68 (95% CI: [0.53–0.82]), F1 score of 0.58 (95% CI: [0.46–0.69]), and AUROC of 0.87 (95% CI: [0.81–0.93]).

**Table 5.** Demographic characteristics of case and control encounters in the 2020 cohort used for FHIR-based AutoML model development, including the overall population and the corresponding training and test datasets *Significant difference between case and control (p <0.05) 1All patients were identified as either male or female 2The last recorded Glasgow coma scale score for the encounter prior to the censored time horizon 3Vasoactive medications include dobutamine, dopamine, epinephrine, norepinephrine, or milrinone 4Sedative-analgesic medications include fentanyl, hydromorphone, midazolam, or morphine Abbreviations: IQR, interquartile range Notes: Continuous variables were compared using Welch’s t-test; categorical variables were compared using Chi-square tests.

|  | Entire 2020 Cohort |  | Train |  | Test |  |
| --- | --- | --- | --- | --- | --- | --- |
|  | Case | Control | Case | Control | Case | Control |
| Size | n=193 | n=1,146 | n=152 | n=874 | n=41 | n=272 |
| Age (months), median (IQR) | 81 (16,178) | 85 (19,173) | 87 (16,177) | 78 (18,168) | 74 (18,186) | 99 (26,182) |
| Female, n (%)<sup>1</sup> | 90(47) | 545(48) | 75(49) | 416(48) | 15(37) | 129(47) |
| Glasgow Coma Scale Score, median (IQR)<sup>2</sup> | 15 (11,15) | 15 (14,15)* | 15 (11,15) | 15 (14,15)* | 14 (10,15) | 15 (12,15)* |
| Mechanical Ventilation, n (%) | 65(34) | 282(25)* | 47(31) | 213(24) | 18(44) | 69(25)* |
| Endotracheal Tube, n(%) | 44(23) | 72(6)* | 29(19) | 51(6)* | 15(37) | 21(8)* |
| Vasoactive Medication, n(%)<sup>3</sup> | 26(13) | 57(5)* | 15(10) | 43(5)* | 11(27) | 14(5)* |
| Sedative-Analgesic Medication, n (%)<sup>4</sup> | 85(44) | 213(19)* | 63(41) | 148(17)* | 22(54) | 65(24)* |

**Table 6.** Performance of FHIR-based AutoML models across training, validation, and test sets for the four experimental settings using a subset of the 2020 cohort. Performance metrics are reported as value (95% confidence interval) for the training and test sets only. Validation performance is reported by AutoML using crossvalidation and does not include AUPRC. Performance on the training and test sets was evaluated via the inference endpoint. The best scores for each metric on the test set are highlighted in bold.

| Feature Set | Baseline |  |  | Complete |  |  | Filtered |  |  | LightGBM |  |  |
| --- | --- | --- | --- | --- | --- | --- | --- | --- | --- | --- | --- | --- |
|  | Train | Validation | Test | Train | Validation | Test | Train | Validation | Test | Train | Validation | Test |
| Accuracy | 0.94<br>(0.92-0.95) | 0.86 | 0.86<br>(0.82-0.90) | 1.00<br>(0.99-1.00) | 0.90 | 0.90<br>(0.87-0.93) | 1.00<br>(1.00-1.00) | 0.91 | **0.90**<br>**(0.87-0.94)** | 0.94<br>(0.92-0.95) | 0.88 | 0.87<br>(0.83-0.91) |
| Precision | 0.74<br>(0.68-0.80) | 0.52 | 0.47<br>(0.35-0.60) | 0.97<br>(0.95-1.00) | 0.68 | 0.63<br>(0.47-0.78) | 1.00<br>(1.00-1.00) | 0.82 | **0.74**<br>**(0.55-0.92)** | 0.74<br>(0.67-0.80) | 0.60 | 0.51<br>(0.38-0.64) |
| Recall | 0.88<br>(0.82-0.93) | 0.57 | 0.63<br>(0.49-0.77) | 1.00<br>(1.00-1.00) | 0.60 | 0.54<br>(0.38-0.69) | 1.00<br>(1.00-1.00) | 0.54 | 0.42<br>(0.26-0.57) | 0.89<br>(0.84-0.93) | 0.67 | **0.68**<br>**(0.53-0.82)** |
| F1 Score | 0.80<br>(0.76-0.85) | 0.54 | 0.54<br>(0.42-0.65) | 0.99<br>(0.97-1.00) | 0.63 | 0.58<br>(0.44-0.70) | 1.00<br>(1.00-1.00) | 0.65 | 0.53<br>(0.37-0.67) | 0.81<br>(0.76-0.85) | 0.63 | **0.58**<br>**(0.46-0.69)** |
| AUROC | 0.97<br>(0.96-0.98) | 0.84 | 0.84<br>(0.77-0.90) | 1.00<br>(1.00-1.00) | 0.88 | 0.85<br>(0.77-0.92) | 1.00<br>(1.00-1.00) | 0.87 | 0.85<br>(0.77-0.91) | 0.98<br>(0.97-0.99) | 0.88 | **0.87**<br>**(0.81-0.93)** |
| AUPRC | 0.88<br>(0.84-0.92) |  | 0.57<br>(0.42-0.71) | 1.00<br>(1.00-1.00) |  | **0.64**<br>**(0.51-0.77)** | 1.00<br>(1.00-1.00) |  | 0.63<br>(0.48-0.76) | 0.90<br>(0.86-0.93) |  | 0.61<br>(0.47-0.75) |

## 5. Discussion

The development of clinical machine learning tools is frequently a boutique process, with each new effort involving customized code for data preparation, model development and evaluation. Some of this complexity is intrinsic to the definitions of features and outcomes relevant to each specific prediction or classification task. Data extraction approaches also impose the need for custom code, as “native” data structures available to researchers may need wrangling to be ready for machine learning. Although commonly used machine learning libraries such as scikit-learn^8^ and Pytorch^10^ support many common machine-learning tasks, workflows for hyperparameter tuning, deep-learning training, and cross-validation are often hand-crafted, differences across implementations.

We envision a future of streamlined and industrialized clinical machine learning model development, with shared infrastructure providing support for common workflows requiring only domain- and problem-specific customization. Just as automobile factories provide assembly lines capable of producing multiple car models with minimal adaptions, clinical machine learning developers will modularize various steps in the process, isolating problem-specific components such as identification of input features, handling of missing data, and definitions of outcomes. These approaches will bring greater rigor, reproducibility, and reusability to model construction without losing the flexibility needed to innovate. In a modularized environment, substituting new approaches for the imputation of missing data or for assessing calibration will be as easy as substituting a new library adhering to specified interfaces. Implementation of these components in a robust computational framework supporting model training, evaluation, and inference will allow for the use of continuous integration/deployment techniques to ensure that models are regularly evaluated and updated.^3,5,21^

Our modeling results demonstrate the robustness of this approach. Models built on the larger dataset extend and improve upon performance demonstrated in our earlier model,^1^ with F1 scores improving from 0.37 to between 0.58 and 0.63 and AUROC improving from 0.81 to 0.88-0.90. Although the larger number of biomarkers (168 vs 44 in the original) may have contributed to this improved performance, the model based on features selected by filtering based on a LightGBM model provided strong performance (F1 0.60, AUROC 0.89) with a smaller overall feature set (525 vs. 584). Strong consistency in the Shapley values across the four feature sets (14 biomarkers occurring in the top 15 for at least 3 of the four models) provide initial validation of the features contributing most to predictions. Results for the smaller FHIR dataset were comparable to those seen in the larger dataset. These results parallel likely deployments, as production FHIR environments might not be configured to provide the years of retrospective data often used to train models.

Our approach takes several concrete steps towards reproducible, production-oriented workflows for clinical machine learning model development. Our use of FHIR for data transport is consistent with US Federal efforts to harmonize data transfer and exchange.^22^ Similarly, our use of AWS FHIR and machine learning services is consistent with recent trends of adoption of cloud services by major medical centers.^23^ As health care organizations continue to build partnerships with cloud providers, clinical decision support models built using these tools might increase alignment with organizational priorities, thus increasing the likelihood of adoption. Cloud provider infrastructure, including standardized libraries, technical support, and the ability to instantiate computational resources on demand, provides powerful and flexible support for modeling work.

Dependence on vendor-specific approaches is a potential downside, as efforts tied too closely to any single vendor’s implementation run the risk of lock-in and decreased opportunities for generalization across institutions. Although the idiosyncrasies of offerings from different vendors will likely prevent direct reuse of code, similarities in the types of offerings provided by the major vendors suggest that translation between systems or construction of appropriate adaptors^24^ will be possible.

Our work is consistent with the continuing trends toward greater transparency and accountability in clinical machine learning efforts. We build upon previous transparency efforts such as model cards^25^ and reporting guidelines,^26^ and draw inspiration from life-cycle models for machine learning, including initial model construction but ongoing monitoring and evaluation.^21,27^ Future extensions to this work will address additional ML operations (MLOps) concerns, including monitoring, retraining, ethics, and regulatory considerations.^3,4^ Our use of FHIR also supports compatibility with emerging tools based on the use of FHIR and USCDI elements to enable federated, cloud-based machine learning.^28^

Limitations include the nature of the FHIR implementation, shortcomings of the definition of neurological morbidity, and the absence of any discussion of potential biases. Although building models directly off FHIR feeds from electronic medical record systems would be preferred as a demonstration of how such models will likely work in practice, many current systems do not directly support FHIR. Our demonstration relied on data extracted from a clinical data warehouse, converted to FHIR format, and stored in an AWS FHIR store. As FHIR-enabled EHR systems become more readily available, this conversion step will likely become unnecessary. The definition of neurological morbidity predicted by our model is a composite of indicators of potential neurological morbidities. There is currently no data showing that intervening on these indicators will lead to improve neurological outcomes. We have also not yet conducted analyses of the impact of health inequities between socioeconomic groups or possible systematic biases in the models.

## 6. Conclusions

Using a combination of AutoML AI and FHIR, we improve upon our previously published results while demonstrating a path toward increased reusability, and generalizability of clinical machine-learning model development processes. By replacing custom machine-learning tools with AutoML pipelines and data wrangling scripts with FHIR, our approach demonstrates the increased transparency and robustness associated with standardized tools. Although the need for custom definitions of specific features and prediction targets will still be required, this framework holds the promise of limiting special-purpose code to discrete steps in the process, relying instead on accepted commodity tools. As regulatory requirements lead to increased adoption of FHIR and related tools by health care providers, these approaches will be increasingly important for supporting clinical adoption of machine-learning tools. Future efforts will focus on further streamlining of this framework.

## 7. Clinical Relevance Statement

Clinical machine-learning algorithms are traditionally developed with *ad hoc* workflows with limited generalizability and reproducibility. We present a standards and tools-based approach that improved performance of a previously published clinical machine-learning model. This use of generalizable processes such as automated machine learning and data transfer standards such as FHIR provides a framework that can be used to create uniform workflows for the development of broadly transferable clinical machine-learning models.

## Supporting information

Complete Feature List

## Data Availability

Data produced in this study are available upon completion of an appropriate data use agreement with the University of Pittsburgh.

## 9. Human Subjects Protections

The study was performed in compliance with the World Medical Association Declaration of Helsinki on Ethical Principles for Medical Research Involving Human Subjects and was reviewed by the University of Pittsburgh Human Research Protection Office (STUDY 20050220).

## 10. Acknowledgements

Qing Liu, and Josh Fleishman from AWS provided valuable assistance with the AWS tools. Dan Ricketts and Uduak S. Ndoh from the University of Pittsburgh helped with data procurement. This research was supported by NIH Grant #R01NS118716.

## Supplemental Materials

**Supplemental Figure 1.**
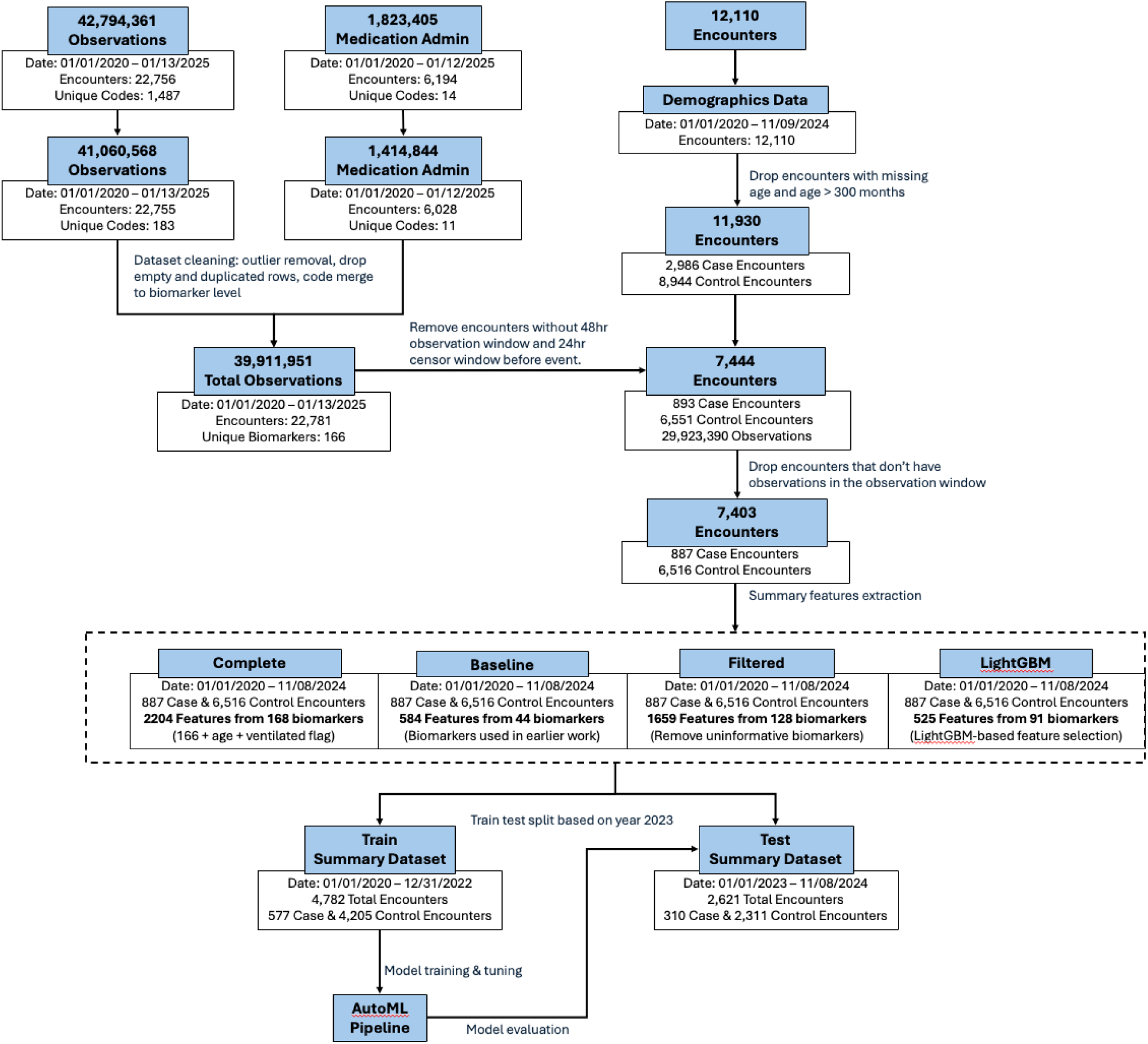
Methodology flowchart illustrating the cohort selection process and corresponding sample sizes at each stage.

**Supplemental Figure 2.**
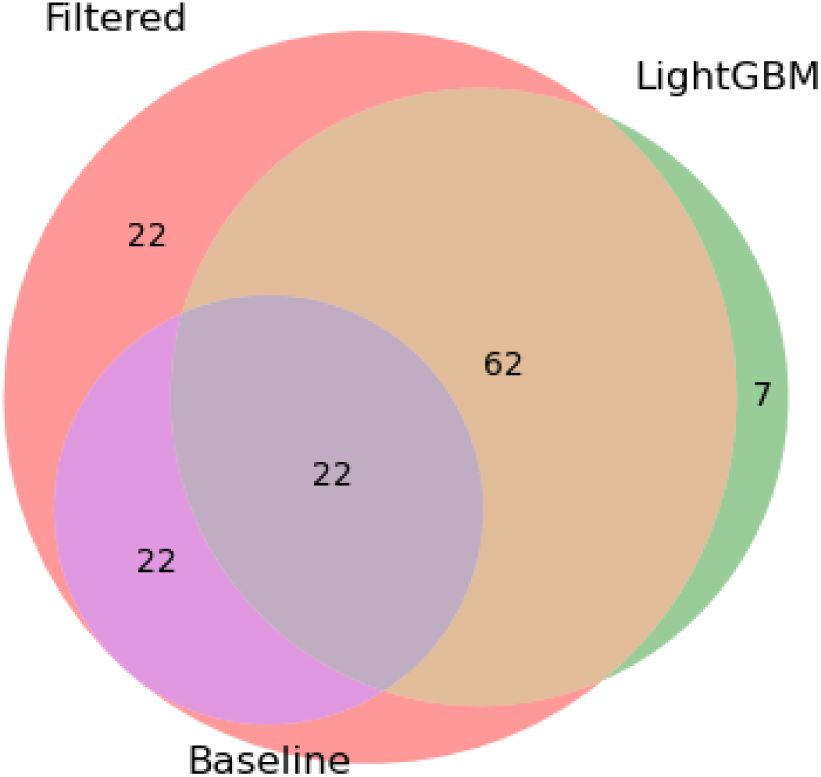
Representation of biomarkers included in the Filtered, LightGBM, and Baseline feature sets. Note that the complete dataset contains the union of these biomarkers (135 biomarkers), plus an additional 33 biomarkers not represented in any of the other feature sets, for a total of 168 biomarkers. The specific biomarkers used in each of the sets can be found in the biomarkers.xlsx supplementary file.

**Supplemental Figure 3.**
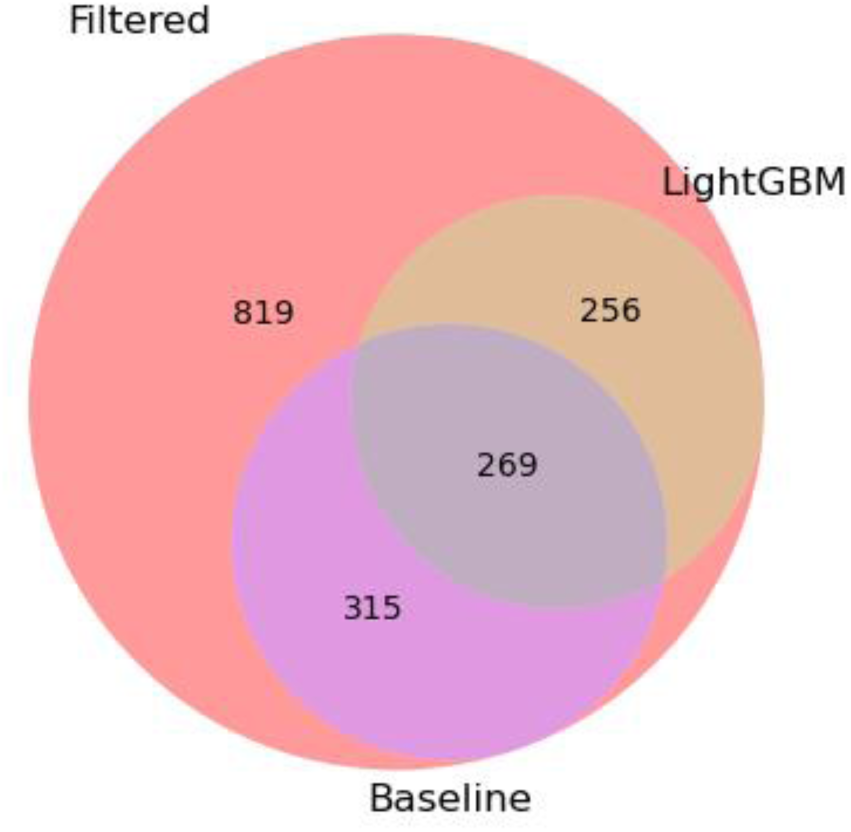
Representation of features in the Filter, LightGBM, and Baseline feature sets. Note that the complete dataset contains the union of these features (1659 features), plus an additional 545 features, for a total of 2,204 features. The specific features used in each of the sets can be found in the features.xlsx supplementary file.

**Supplemental Figure 4.**
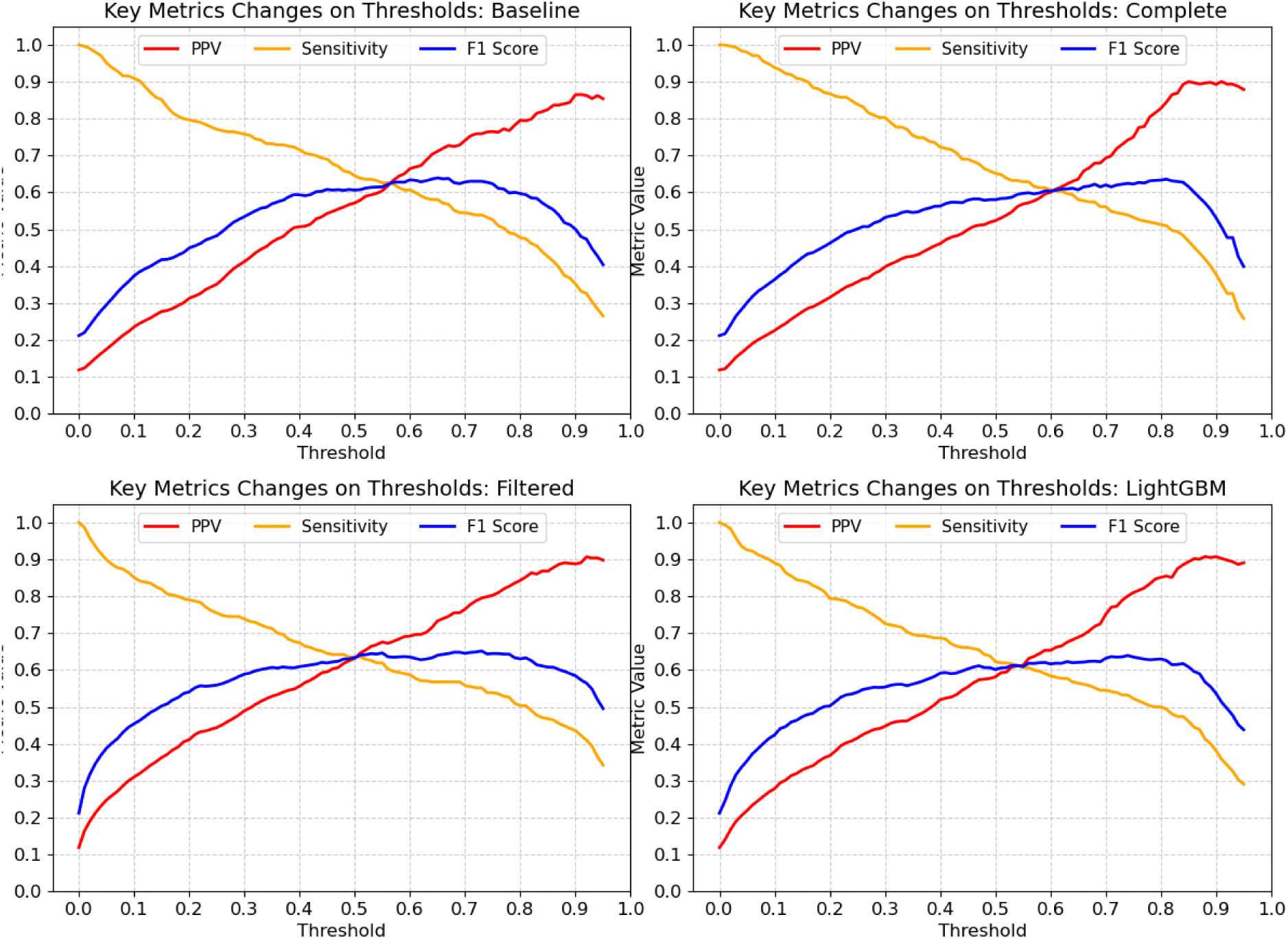
Performance metric curves showing changes in positive predictive value (PPV), sensitivity, and F1 score across decision thresholds for each of the four feature sets (Baseline, Complete, Filtered, and LightGBM) , evaluated on the test set.

**Supplemental Figure 5.**
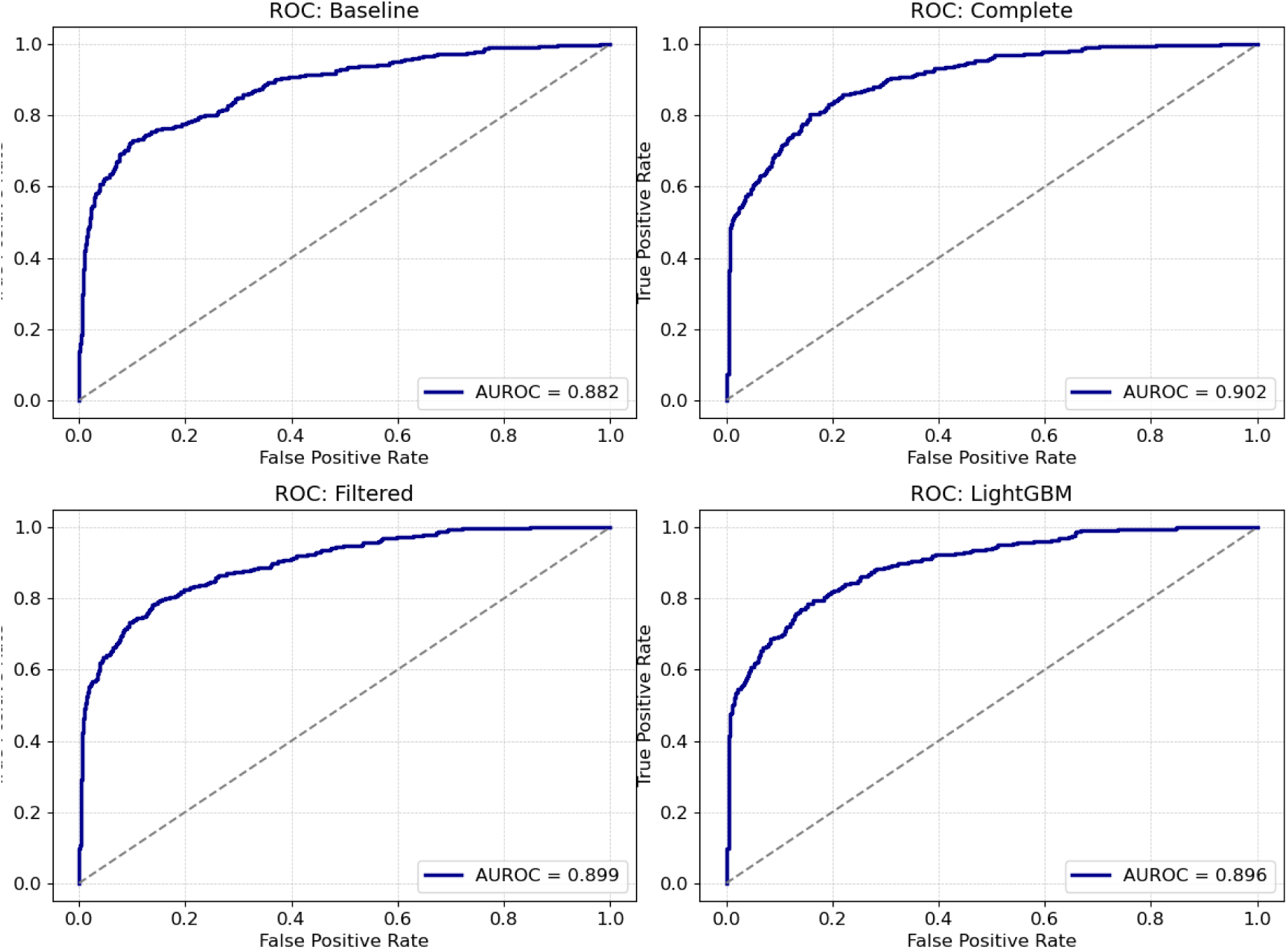
ROC curves for the AutoML models using each of the four feature sets (Baseline, Complete, Filtered, and LightGBM), evaluated on the test set.

**Supplemental Figure 6.**
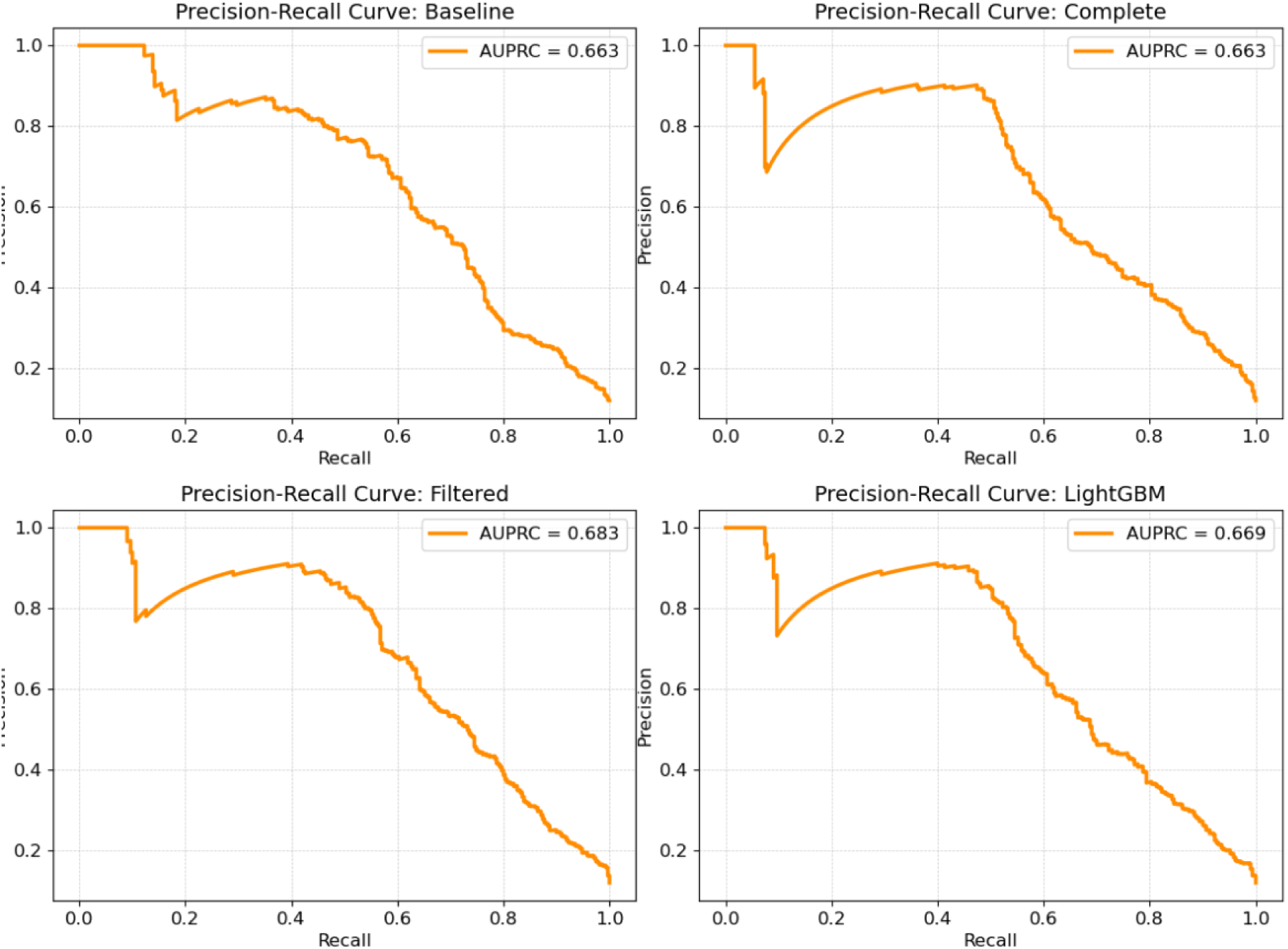
Precision-recall (PR) curves for the AutoML models using each of the four feature sets (Baseline, Complete, Filtered, and LightGBM), evaluated on the test set.

**Supplemental Figure 7.**
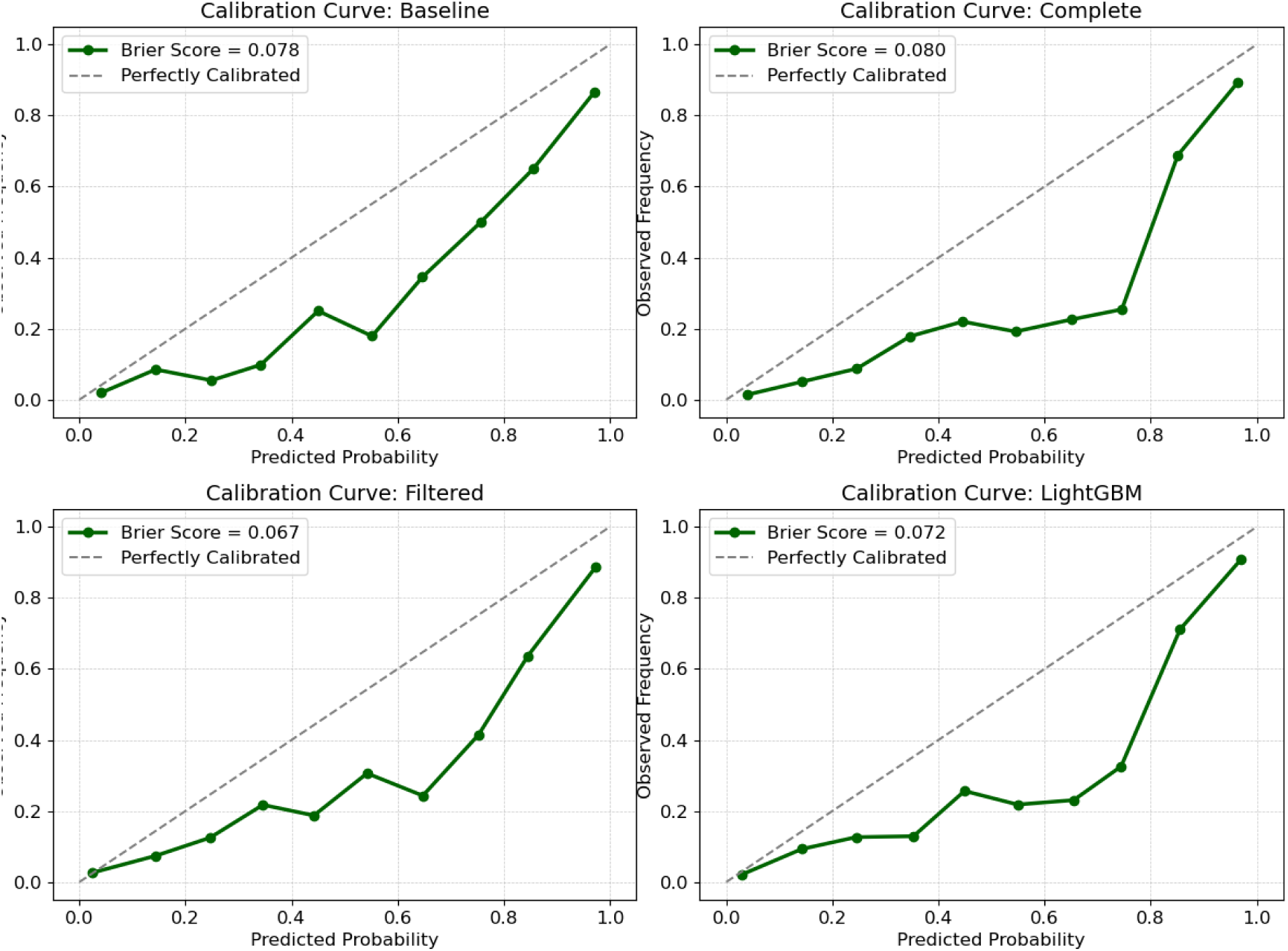
Calibration curves for the AutoML models using each of the four feature sets (Baseline, Complete, Filtered, and LightGBM), evaluated on the test set.

**Supplemental Table 1.** Data curation steps for each numerical data element.

| Biomarker_Name | Type | Min | Max | Action |
| --- | --- | --- | --- | --- |
| Packed erythrocytes given [Volume] | Numerical | 0 | 2800 | Truncate |
| Alanine aminotransferase [Enzymatic activity/volume] in Serum or Plasma | Numerical | 6 | 12000 | Truncate |
| Albumin [Mass/volume] in Serum or Plasma | Numerical | 0 | 7 | Truncate |
| Volume of Stool | Numerical | 0 | 2400 | Truncate |
| Calcium [Mass/volume] in Serum or Plasma | Numerical | 2 | 30 | Truncate |
| Magnesium [Mass/volume] in Serum or Plasma | Numerical | 0.3 | 13 | Truncate |
| Aspartate aminotransferase [Enzymatic activity/volume] in Serum or Plasma | Numerical | 0 | 20000 | Truncate |
| Bilirubin.direct [Mass/volume] in Serum or Plasma | Numerical | 0.1 | 60 | Truncate |
| Bilirubin.total [Mass/volume] in Serum or Plasma | Numerical | 0.1 | 60 | Truncate |
| Carbon dioxide [Partial pressure] in Exhaled gas --at end expiration | Numerical | 0 | 150 | Truncate |
| Calcium.ionized [Moles/volume] in Blood | Numerical | 0.2 | 3 | Truncate |
| Humpty Dumpty fall risk assessment score (observable entity) | Numerical | 7 | 23 | Discard |
| Positive end expiratory pressure setting Ventilator | Numerical | 0 | 40 | Discard |
| Tidal volume setting Ventilator | Numerical | 10 | 1000 | Discard |
| Carbon dioxide [Partial pressure] in Venous blood | Numerical | 5 | 150 | Truncate |
| Hemoglobin [Mass/volume] in Blood by calculation | Numerical | 0 | 30 | Discard |
| Gamma glutamyl transferase [Enzymatic activity/volume] in Serum or Plasma | Numerical | 3 | 4000 | Truncate |
| Osmolality of Serum or Plasma | Numerical | 50 | 500 | Truncate |
| Oxygen [Partial pressure] in Arterial blood | Numerical | 12 | 725 | Discard |
| Oxygen [Partial pressure] in Venous blood | Numerical | 10 | 600 | Discard |
| pH of Urine | Numerical | 4 | 8 | Discard |
| Phosphate [Mass/volume] in Serum or Plasma | Numerical | 0.1 | 18 | Truncate |
| Protein [Mass/volume] in Serum or Plasma | Numerical | 1 | 10 | Truncate |
| Inhaled oxygen concentration | Numerical | 18 | 100 | Discard |
| Inhaled oxygen flow rate | Numerical | 0 | 100 | Discard |
| Fibrinogen [Mass/volume] in Platelet poor plasma by Coagulation assay | Numerical | 19 | 1800 | Truncate |
| Platelet mean volume [Entitic volume] in Blood by Automated count | Numerical | 3 | 15 | Truncate |
| Anion gap in Serum or Plasma | Numerical | 0 | 40 | Truncate |
| Creatinine renal clearance/1.73 sq M.predicted by Cockcroft-Gault formula, BSA formula | Numerical | 0 | 200 | Discard |
| Pain severity total [Score] FLACC | Numerical | 0 | 10 | Discard |
| Body mass index (BMI) [Ratio] | Numerical | 10 | 50 | Discard |
| Vancomycin [Mass/volume] in Serum or Plasma --trough | Numerical | 0 | 65 | Truncate |
| Hematocrit [Volume Fraction] of Blood by Automated count | Numerical | 5 | 75 | Truncate |
| Specific gravity of Urine by Test strip | Numerical | 0.8 | 1.2 | Discard |
| Monocytes/100 leukocytes in Blood by Automated count | Numerical | 0 | 60 | Truncate |
| Prothrombin time (PT) in Blood by Coagulation assay | Numerical | 5 | 120 | Truncate |
| Intracranial pressure (ICP) | Numerical | -40 | 120 | Truncate |
| Central venous pressure (CVP) | Numerical | -12 | 60 | Truncate |
| Cerebral perfusion pressure | Numerical | 0 | 200 | Truncate |
| Alkaline phosphatase [Enzymatic activity/volume] in Serum or Plasma | Numerical | 10 | 2400 | Truncate |
| Beta hydroxybutyrate [Moles/volume] in Serum or Plasma | Numerical | 0 | 14 | Truncate |
| Basophils [# /volume] in Blood by Automated count | Numerical | 0 | 10 | Truncate |
| Basophils/100 leukocytes in Blood by Automated count | Numerical | 0 | 55 | Truncate |
| Eosinophils [# /volume] in Blood by Automated count | Numerical | 0 | 100 | Discard |
| Eosinophils/100 leukocytes in Blood by Automated count | Numerical | 0 | 100 | Discard |
| Lymphocytes [# /volume] in Blood by Automated count | Numerical | 0 | 100 | Discard |
| Variant lymphocytes/100 leukocytes in Blood by Manual count | Numerical | 0 | 80 | Truncate |
| Lymphocytes/100 leukocytes in Blood by Automated count | Numerical | 0 | 100 | Discard |
| Metamyelocytes [# /volume] in Blood by Manual count | Numerical | 0 | 65 | Truncate |
| Metamyelocytes/100 leukocytes in Blood by Manual count | Numerical | 0 | 65 | Truncate |
| Tacrolimus [Mass/volume] in Blood by LC/MS/MS | Numerical | 0 | 200 | Truncate |
| Monocytes [# /volume] in Blood by Automated count | Numerical | 0 | 60 | Truncate |
| Myelocytes [# /volume] in Blood by Manual count | Numerical | 0 | 30 | Truncate |
| Myelocytes/100 leukocytes in Blood by Manual count | Numerical | 0 | 30 | Truncate |
| Neutrophils [# /volume] in Blood by Automated count | Numerical | 0 | 400 | Truncate |
| Tidal volume expired Respiratory system airway --on ventilator | Numerical | 0 | 2000 | Discard |
| Band form neutrophils [# /volume] in Blood by Manual count | Numerical | 0 | 70 | Truncate |
| Band form neutrophils/100 leukocytes in Blood by Manual count | Numerical | 0 | 70 | Truncate |
| Gestational age--at birth | Numerical | 154 | 308 | Truncate |
| Neutrophils/100 leukocytes in Blood by Automated count | Numerical | 0 | 100 | Discard |
| MCH [Entitic mass] by Automated count | Numerical | 0 | 75 | Discard |
| MCHC [Mass/volume] by Automated count | Numerical | 28 | 325 | Truncate |
| MCV [Entitic volume] by Automated count | Numerical | 40 | 150 | Truncate |
| Erythrocyte distribution width [Ratio] by Automated count | Numerical | 10 | 70 | Truncate |
| Erythrocytes [# /volume] in Blood by Automated count | Numerical | 0 | 10 | Truncate |
| Body weight --used for drug calculation | Numerical | 0 | 300 | Discard |
| Body surface area | Numerical | 0.1 | 5 | Discard |
| Body height | Numerical | 25 | 250 | Truncate |
| Base deficit | Numerical | -30 | 0 | Discard |
| Base excess | Numerical | 0 | 30 | Discard |
| Bicarbonate | Numerical | 0 | 80 | Discard |
| Blood urea nitrogen | Numerical | 1 | 300 | Discard |
| C-Reactive Protein | Numerical | 0 | 100 | Discard |
| Chloride | Numerical | 60 | 190 | Discard |
| Creatinine | Numerical | 0.1 | 25 | Discard |
| DBP | Numerical | 0 | 200 | Discard |
| Glucose | Numerical | 0 | 2000 | Discard |
| Hemoglobin | Numerical | 0 | 30 | Discard |
| INR | Numerical | 0 | 25 | Truncate |
| Lactate | Numerical | 0 | 30 | Discard |
| MBP | Numerical | 0 | 160 | Discard |
| PTT | Numerical | 0 | 250 | Truncate |
| Peds Coma Score | Numerical | 3 | 15 | Discard |
| Platelets | Numerical | 0 | 5000 | Discard |
| Potassium | Numerical | 0.05 | 12 | Discard |
| Procalcitonin | Numerical | 0 | 250 | Discard |
| Pulse | Numerical | 0 | 350 | Discard |
| Respiratory Rate | Numerical | 0 | 150 | Discard |
| SBP | Numerical | 0 | 300 | Discard |
| Sodium | Numerical | 80 | 215 | Discard |
| SpO2 | Numerical | 0 | 100 | Discard |
| Temperature | Numerical | 0 | 46 | Discard |
| Weight | Numerical | 0 | 300 | Discard |
| White blood cell count | Numerical | 0 | 1300 | Truncate |
| Pco2 | Numerical | 5 | 150 | Truncate |
| pH | Numerical | 6 | 8 | Truncate |

**Supplemental Table 2:** Hyperparameter configurations for models trained in each experiment using the entire study and the 2020 subset of the cohort. Training configuration parameters: _kfold: Number of folds used for cross-validation during model training; _tuning_objective_metric: The metric that Autopilot used to tune the model. eval_metric: Evaluation metric used to evaluate model performance during training; objective: Specifies the learning task and corresponding loss function (“binary:logistic” for binary classification). Regularization parameters: alpha: L1 regularization term (larger alpha, sparsity increases); lambda: L2 regularization term (larger lambda, model generalization improves). Model parameters: subsample: Fraction of training samples used per tree (like dropout for boosting); colsample_bytree: Fraction of features randomly selected for each tree (prevents overfitting); max_depth: Maximum depth of a tree (higher = more complexity, risk of overfitting); min_child_weight: Minimum sum of instance weight (hessian) needed in a child (higher = more conservative tree); eta: Learning rate (smaller = slower learning, better generalization); gamma: Minimum loss reduction to make a split (higher values = more conservative); num_round: Number of boosting rounds/trees to build; scale_pos_weight: Controls balance of positive and negative weights (useful for imbalanced datasets).

| **Entire Cohort** |  |  |  |  |
| --- | --- | --- | --- | --- |
| Hyperparameters | Baseline | Complete | Filtered | LightGBM |
| `kfold` | 5 |  |  |  |
| `_tuning_objective_metric` | validation:f1_binary |  |  |  |
| `eval_metric` | accuracy, f1_binary, balanced accuracy, precision, recall, logloss |  |  |  |
| `objective` | binary:logistic |  |  |  |
| `alpha` | 2.674e-6 | 0.622 | 0.053 | 1.241 |
| `lambda` | 0.012 | 0.000 | 1.222e-5 | 1.952e-6 |
| `subsample` | 0.624 | 0.842 | 0.735 | 0.650 |
| `colsample_bytree` | 0.311 | 0.665 | 0.338 | 0.492 |
| `max_depth` | 2 | 3 | 2 | 6 |
| `min_child_weight` | 27.854 | 1.673e-6 | 1.044 | 1.014e-6 |
| `eta` | 0.085 | 0.027 | 0.079 | 0.074 |
| `gamma` | 0.207 | 1.286e-5 | 1.517 | 8.699 |
| `num_round` | 535 | 521 | 889 | 703 |
| `scale_pos_weight` | 7.288 | 7.288 | 7.288 | 7.288 |
| **2020 Subset** |  |  |  |  |
| `kfold` | 5 |  |  |  |
| `_tuning_objective_metric` | validation:f1_binary |  |  |  |
| `eval_metric` | accuracy, f1_binary, balanced accuracy, precision, recall, logloss |  |  |  |
| `objective` | binary:logistic |  |  |  |
| `alpha` | 0.026 | 0.000 | 5.033e-6 | 0.001 |
| `lambda` | 0.125 | 1.052e-5 | 0.070 | 9.205e-6 |
| `subsample` | 0.921 | 0.779 | 0.865 | 0.716 |
| `colsample_bytree` | 0.449 | 0.720 | 0.811 | 0.336 |
| `max_depth` | 3 | 2 | 3 | 5 |
| `min_child_weight` | 29.037 | 9.728e-5 | 5.378e-5 | 3.760e-6 |
| `eta` | 0.004 | 0.132 | 0.156 | 0.108 |
| `gamma` | 0.009 | 4.536 | 1.381 | 36.630 |
| `num_round` | 716 | 86 | 317 | 664 |
| `scale_pos_weight` | 5.75 | 5.75 | 5.75 | 5.75 |

**Supplemental Table 3.** Summary statistics for 96 numerical biomarkers used in the analysis (N = 7403 encounters). For each biomarker, the table reports the number of encounters with at least one recorded value, the number and percentage of encounters missing values, and the median, 25th percentile (Q25), and 75th percentile (Q75) of the recorded values.

| Biomarker_Name | Enc_HasBiomarker<br>(N) | Enc_NoBiomarker<br>(N) | Enc_NoBiomarker<br>(%) | Median | Q25 | Q75 |
| --- | --- | --- | --- | --- | --- | --- |
| Packed erythrocytes given [Volume] | 890 | 6513 | 88.0 | 77.5 | 31.3 | 120.5 |
| Alanine aminotransferase [Enzymatic activity/volume] in Serum or Plasma | 3316 | 4087 | 55.2 | 48 | 26 | 116 |
| Albumin [Mass/volume] in Serum or Plasma | 3605 | 3798 | 51.3 | 3 | 2.6 | 3.4 |
| Volume of Stool | 2351 | 5052 | 68.2 | 40 | 8.5 | 113 |
| Calcium [Mass/volume] in Serum or Plasma | 7143 | 260 | 3.5 | 8.7 | 8.2 | 9.2 |
| Magnesium [Mass/volume] in Serum or Plasma | 6864 | 539 | 7.3 | 2 | 1.8 | 2.3 |
| Aspartate aminotransferase [Enzymatic activity/volume] in Serum or Plasma | 3286 | 4117 | 55.6 | 42 | 25 | 99 |
| Bilirubin.direct [Mass/volume] in Serum or Plasma | 2689 | 4714 | 63.7 | 0.3 | 0.1 | 0.9 |
| Bilirubin.total [Mass/volume] in Serum or Plasma | 3281 | 4122 | 55.7 | 0.6 | 0.3 | 1.6 |
| Carbon dioxide [Partial pressure] in Exhaled gas --at end expiration | 1829 | 5574 | 75.3 | 37 | 34 | 41 |
| Calcium.ionized [Moles/volume] in Blood | 4408 | 2995 | 40.5 | 1.3 | 1.2 | 1.4 |
| Humpty Dumpty fall risk assessment score (observable entity) | 7200 | 203 | 2.7 | 14 | 11 | 16 |
| Positive end expiratory pressure setting Ventilator | 2316 | 5087 | 68.7 | 7 | 6 | 9 |
| Tidal volume setting Ventilator | 446 | 6957 | 94.0 | 200 | 75 | 350 |
| Carbon dioxide [Partial pressure] in Venous blood | 3976 | 3427 | 46.3 | 45 | 36 | 53 |
| Hemoglobin [Mass/volume] in Blood by calculation | 4363 | 3040 | 41.1 | 10.2 | 8.8 | 11.9 |
| Gamma glutamyl transferase [Enzymatic activity/volume] in Serum or Plasma | 994 | 6409 | 86.6 | 96 | 43 | 217 |
| Osmolality of Serum or Plasma | 1077 | 6326 | 85.5 | 302 | 292 | 316 |
| Oxygen [Partial pressure] in Arterial blood | 1286 | 6117 | 82.6 | 102 | 77 | 141 |
| Oxygen [Partial pressure] in Venous blood | 3969 | 3434 | 46.4 | 43 | 36 | 53 |
| pH of Urine | 1078 | 6325 | 85.4 | 6 | 6 | 7 |
| Phosphate [Mass/volume] in Serum or Plasma | 6929 | 474 | 6.4 | 4 | 3.2 | 4.8 |
| Protein [Mass/volume] in Serum or Plasma | 3256 | 4147 | 56.0 | 5.7 | 5 | 6.5 |
| Inhaled oxygen concentration | 1578 | 5825 | 78.7 | 38 | 28.8 | 58 |
| Inhaled oxygen flow rate | 5094 | 2309 | 31.2 | 3 | 1 | 8 |
| Fibrinogen [Mass/volume] in Platelet poor plasma by Coagulation assay | 1506 | 5897 | 79.7 | 237 | 158 | 346.8 |
| Platelet mean volume [Entitic volume] in Blood by Automated count | 6227 | 1176 | 15.9 | 8.3 | 7.6 | 9.2 |
| Anion gap in Serum or Plasma | 7151 | 252 | 3.4 | 7 | 5 | 9 |
| Creatinine renal clearance/1.73 sq M.predicted by Cockcroft-Gault formula, BSA formula | 2372 | 5031 | 68.0 | 98.6 | 58.2 | 138.3 |
| Pain severity total [Score] FLACC | 6764 | 639 | 8.6 | 0 | 0 | 0 |
| Body mass index (BMI) [Ratio] | 5225 | 2178 | 29.4 | 17.4 | 15.1 | 20.9 |
| Vancomycin [Mass/volume] in Serum or Plasma --trough | 2068 | 5335 | 72.1 | 11.7 | 8.3 | 15.7 |
| Hematocrit [Volume Fraction] of Blood by Automated count | 6886 | 517 | 7.0 | 29.5 | 25.6 | 34 |
| Specific gravity of Urine by Test strip | 3335 | 4068 | 55.0 | 1.02 | 1.0 | 1.0 |
| Monocytes/100 leukocytes in Blood by Automated count | 6333 | 1070 | 14.5 | 8 | 5 | 11 |
| Prothrombin time (PT) in Blood by Coagulation assay | 2854 | 4549 | 61.5 | 16.3 | 14.7 | 19.5 |
| Intracranial pressure (ICP) | 42 | 7361 | 99.4 | 7 | 4 | 10 |
| Central venous pressure (CVP) | 625 | 6778 | 91.6 | 8 | 5 | 12 |
| Cerebral perfusion pressure | 18 | 7385 | 99.8 | 71 | 60 | 82 |
| Alkaline phosphatase [Enzymatic activity/volume] in Serum or Plasma | 3283 | 4120 | 55.7 | 149 | 96 | 240 |
| Beta hydroxybutyrate [Moles/volume] in Serum or Plasma | 952 | 6451 | 87.1 | 1.3 | 0.4 | 3.5 |
| Basophils [# /volume] in Blood by Automated count | 5824 | 1579 | 21.3 | 0.0 | 0.0 | 0.1 |
| Basophils/100 leukocytes in Blood by Automated count | 5824 | 1579 | 21.3 | 0 | 0 | 1 |
| Eosinophils [# /volume] in Blood by Automated count | 5957 | 1446 | 19.5 | 0.1 | 0.0 | 0.3 |
| Eosinophils/100 leukocytes in Blood by Automated count | 5957 | 1446 | 19.5 | 1 | 0 | 3 |
| Lymphocytes [# /volume] in Blood by Automated count | 6343 | 1060 | 14.3 | 1.3 | 0.6 | 2.5 |
| Variant lymphocytes/100 leukocytes in Blood by Manual count | 2477 | 4926 | 66.5 | 2 | 1 | 3 |
| Lymphocytes/100 leukocytes in Blood by Automated count | 6343 | 1060 | 14.3 | 16 | 7 | 30 |
| Metamyelocytes [# /volume] in Blood by Manual count | 1741 | 5662 | 76.48 | 0.2 | 0.1 | 0.4 |
| Metamyelocytes/100 leukocytes in Blood by Manual count | 1740 | 5663 | 76.5 | 2 | 1 | 3 |
| Tacrolimus [Mass/volume] in Blood by LC/MS/MS | 419 | 6984 | 94.34 | 7.8 | 5.1 | 11 |
| Monocytes [# /volume] in Blood by Automated count | 6333 | 1070 | 14.45 | 0.7 | 0.3 | 1.1 |
| Myelocytes [# /volume] in Blood by Manual count | 1634 | 5769 | 77.93 | 0.2 | 0.1 | 0.4 |
| Myelocytes/100 leukocytes in Blood by Manual count | 1633 | 5770 | 77.94 | 2 | 1 | 3 |
| Neutrophils [# /volume] in Blood by Automated count | 6344 | 1059 | 14.31 | 5.7 | 3.1 | 9.8 |
| Tidal volume expired Respiratory system airway --on ventilator | 3165 | 4238 | 57.25 | 137 | 72 | 271 |
| Band form neutrophils [# /volume] in Blood by Manual count | 2106 | 5297 | 71.55 | 0.2 | 0.1 | 0.5 |
| Band form neutrophils/100 leukocytes in Blood by Manual count | 2105 | 5298 | 71.57 | 2 | 1 | 4 |
| Gestational age--at birth | 3153 | 4250 | 57.41 | 258 | 231 | 273 |
| Neutrophils/100 leukocytes in Blood by Automated count | 6344 | 1059 | 14.31 | 70 | 54 | 83 |
| MCH [Entitic mass] by Automated count | 6375 | 1028 | 13.89 | 29.1 | 27.6 | 30.4 |
| MCHC [Mass/volume] by Automated count | 6375 | 1028 | 13.89 | 33.8 | 33 | 34.5 |
| MCV [Entitic volume] by Automated count | 6375 | 1028 | 13.89 | 85.9 | 82.1 | 89.3 |
| Erythrocyte distribution width [Ratio] by Automated count | 6373 | 1030 | 13.91 | 15.7 | 14.2 | 17.4 |
| Erythrocytes [# /volume] in Blood by Automated count | 6375 | 1028 | 13.89 | 3.4 | 3.0 | 3.9 |
| Body weight --used for drug calculation | 5771 | 1632 | 22.05 | 13.3 | 5.4 | 37.9 |
| Body surface area | 4614 | 2789 | 37.67 | 0.5 | 0.3 | 1.2 |
| Body height | 5603 | 1800 | 24.31 | 108 | 68.5 | 148 |
| Base deficit | 3647 | 3756 | 50.74 | -4 | -10 | -2 |
| Base excess | 2589 | 4814 | 65.03 | 4 | 2 | 8 |
| Bicarbonate | 7198 | 205 | 2.77 | 25 | 22 | 29 |
| Blood urea nitrogen | 7149 | 254 | 3.43 | 13 | 8 | 22 |
| C-Reactive Protein | 5003 | 2400 | 32.42 | 2.2 | 0.7 | 6.8 |
| Chloride | 7153 | 250 | 3.38 | 109 | 105 | 113 |
| Creatinine | 7148 | 255 | 3.44 | 0.4 | 0.2 | 0.7 |
| DBP | 7392 | 11 | 0.15 | 58 | 49 | 68 |
| Glucose | 7169 | 234 | 3.16 | 115 | 96 | 157 |
| Hemoglobin | 6379 | 1024 | 13.83 | 9.8 | 8.6 | 11.3 |
| INR | 2854 | 4549 | 61.45 | 1.3 | 1.2 | 1.7 |
| Lactate | 1635 | 5768 | 77.91 | 1.5 | 1 | 2.5 |
| MBP | 7390 | 13 | 0.18 | 74 | 64 | 85 |
| PTT | 2821 | 4582 | 61.89 | 39 | 31 | 60 |
| Peds Coma Score | 7402 | 1 | 0.01 | 13 | 9 | 15 |
| Platelets | 6361 | 1042 | 14.08 | 184 | 92 | 299 |
| Potassium | 7153 | 250 | 3.38 | 3.8 | 3.3 | 4.3 |
| Procalcitonin | 4721 | 2682 | 36.23 | 0.4 | 0.1 | 2.4 |
| Pulse | 7392 | 11 | 0.15 | 112 | 91 | 132 |
| Respiratory Rate | 7392 | 11 | 0.15 | 27 | 20 | 38 |
| SBP | 7392 | 11 | 0.15 | 104 | 92 | 117 |
| Sodium | 7206 | 197 | 2.66 | 140 | 137 | 143 |
| SpO2 | 7392 | 11 | 0.15 | 98 | 97 | 100 |
| Temperature | 7392 | 11 | 0.15 | 36.7 | 36.4 | 37 |
| Weight | 6867 | 536 | 7.24 | 14.8 | 7.2 | 37.2 |
| White blood cell count | 6373 | 1030 | 13.91 | 8.7 | 5.2 | 13.4 |
| Pco2 | 2252 | 5151 | 69.58 | 44 | 38 | 53 |
| pH | 4542 | 2861 | 38.65 | 7.4 | 7.3 | 7.4 |

**Supplemental Table 4.**
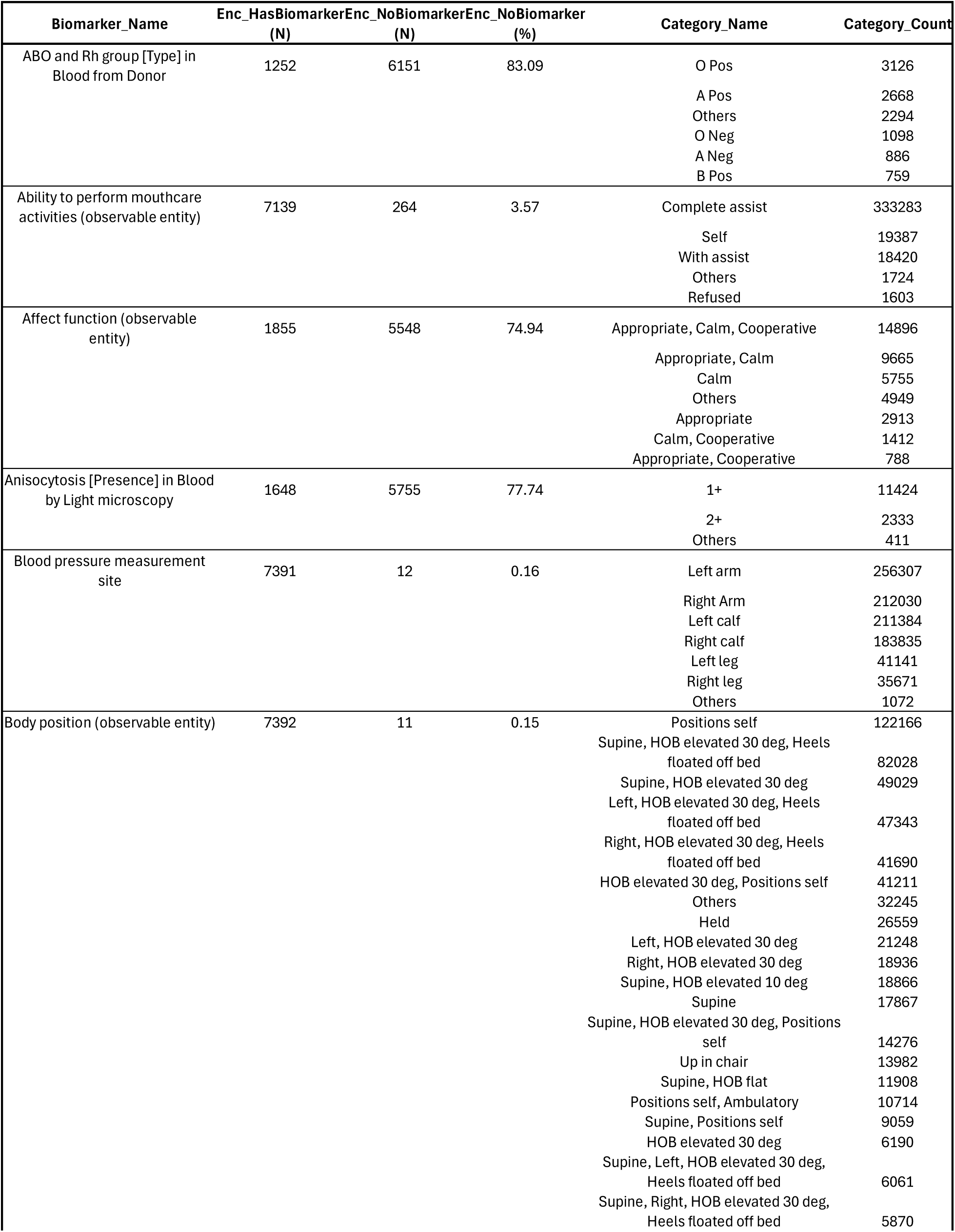
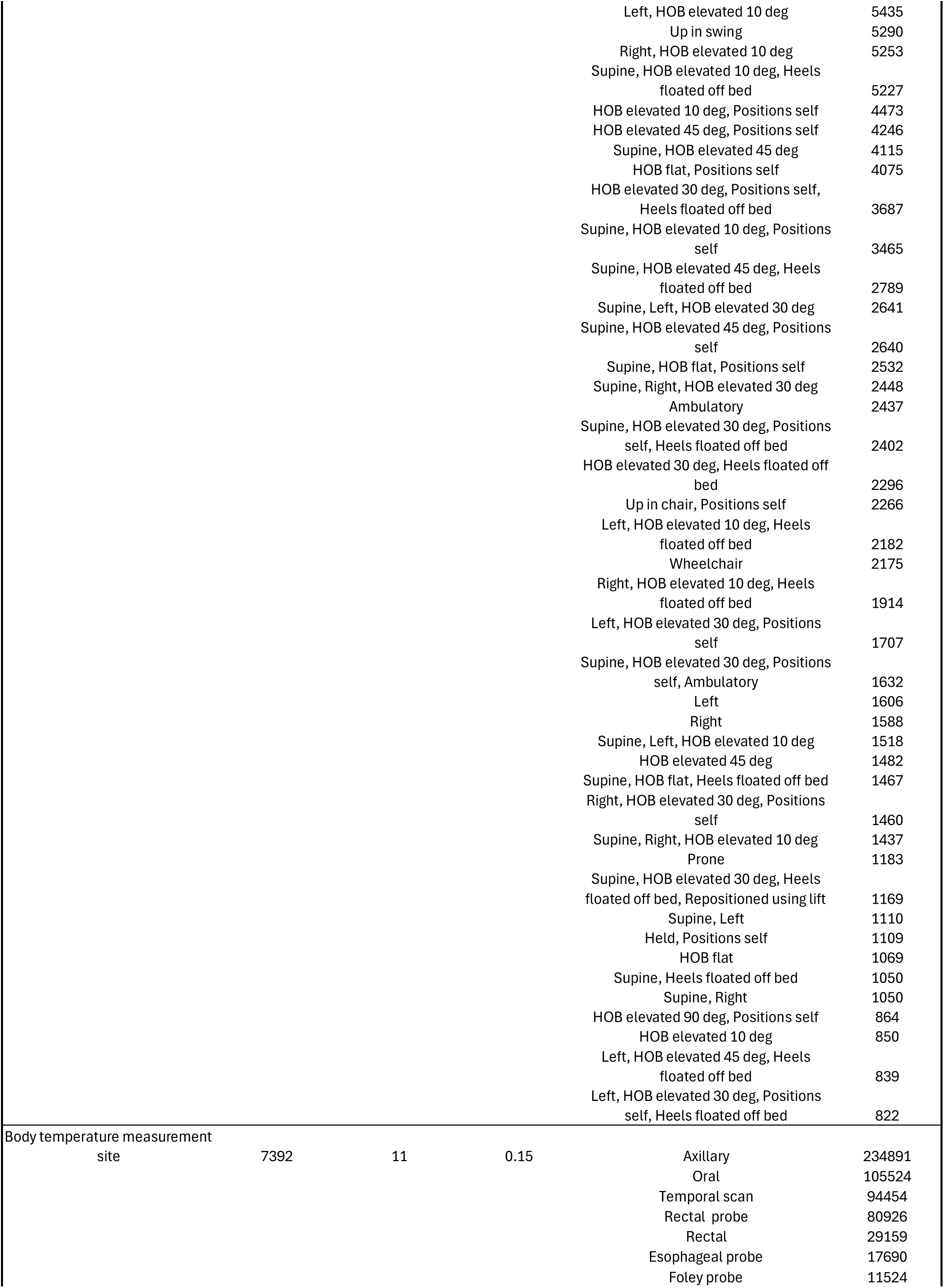
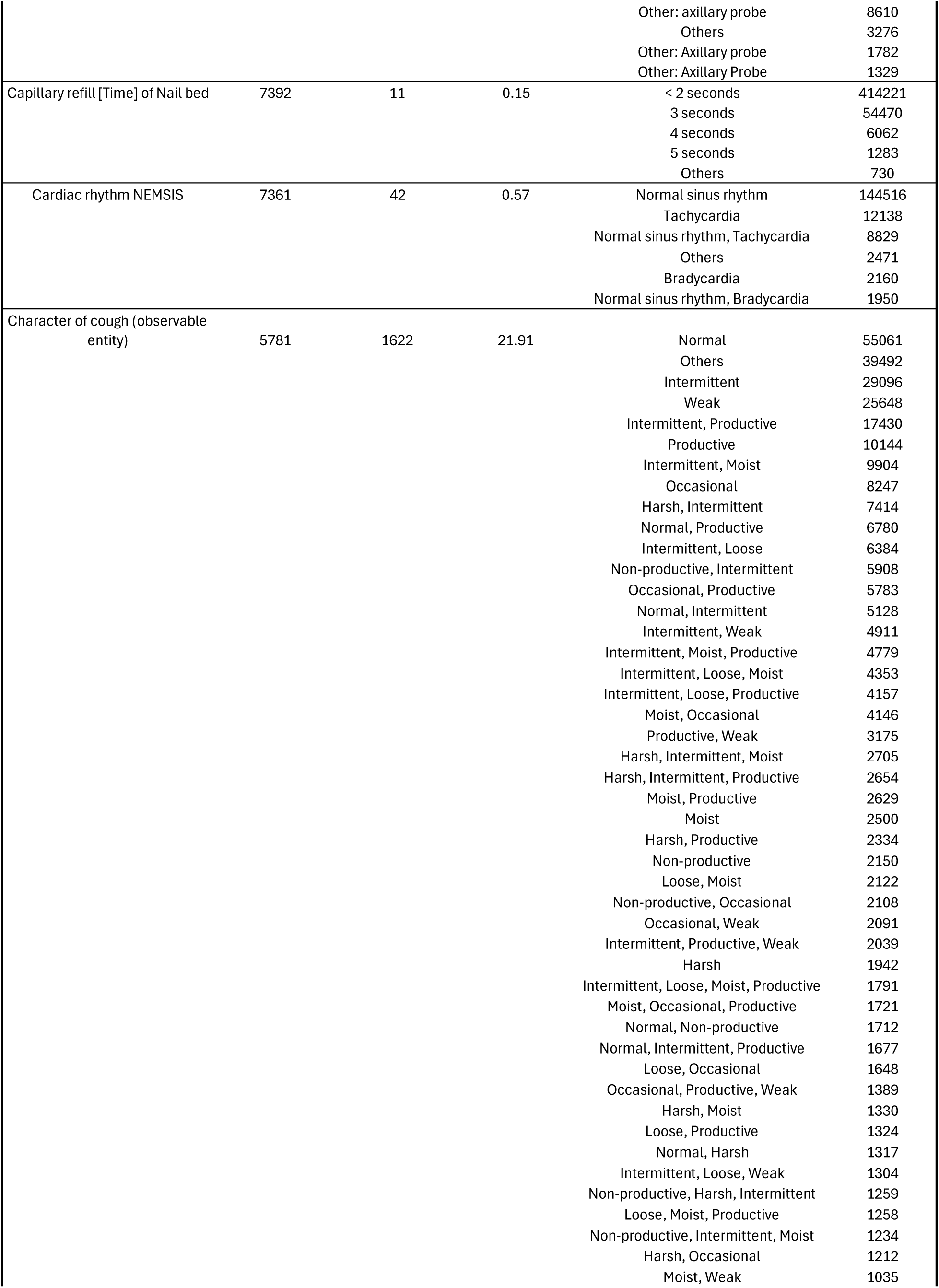
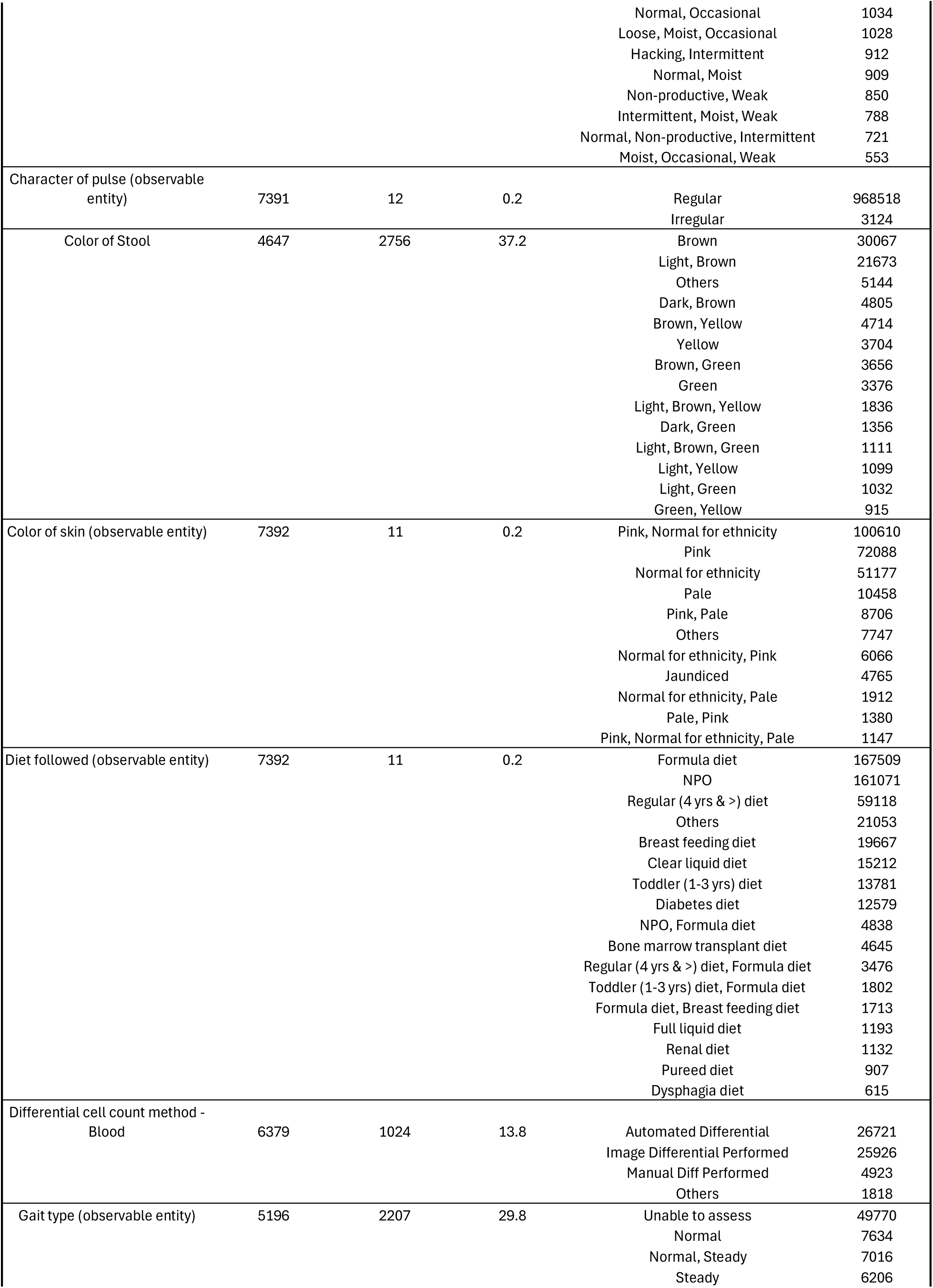
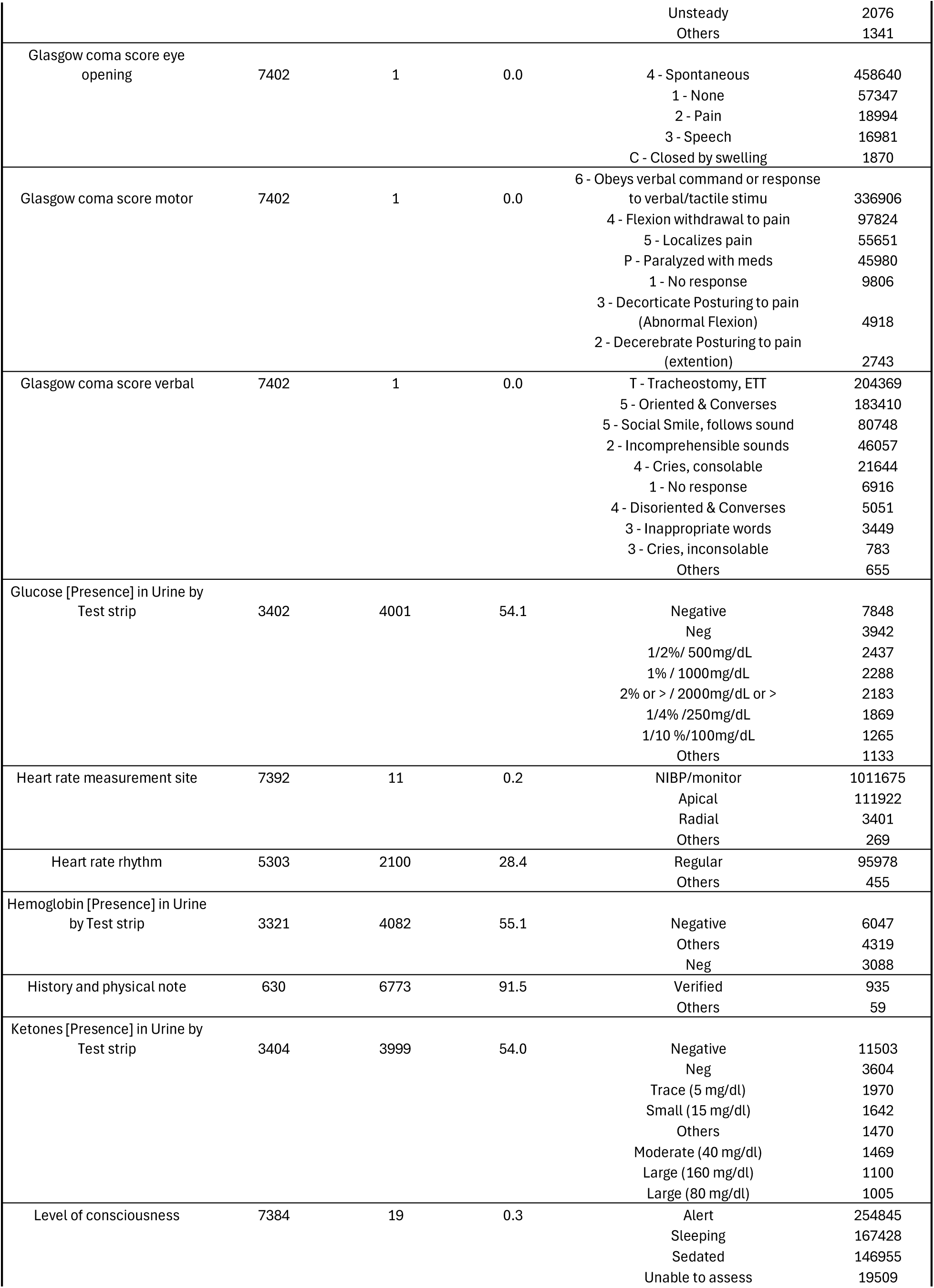
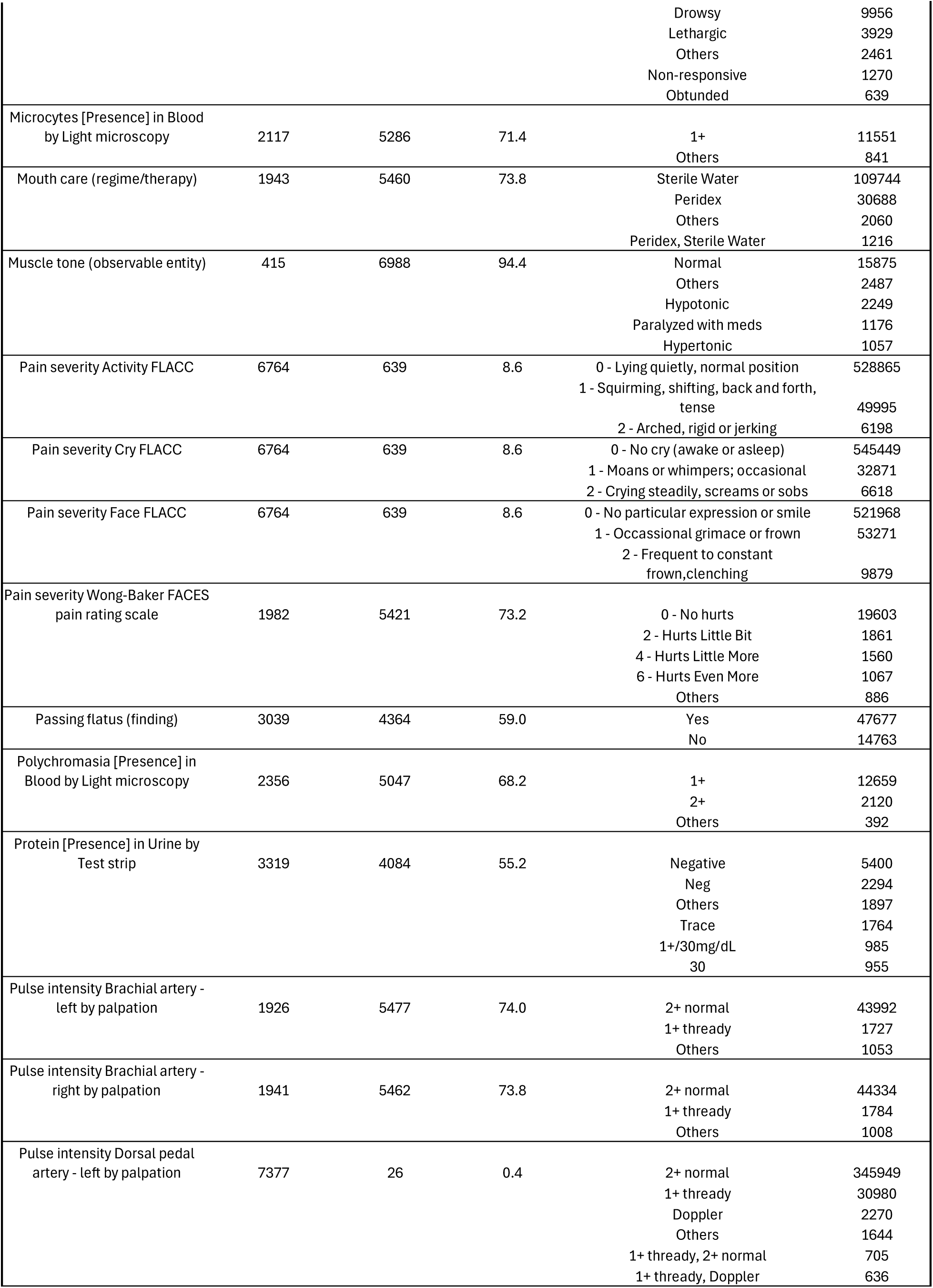
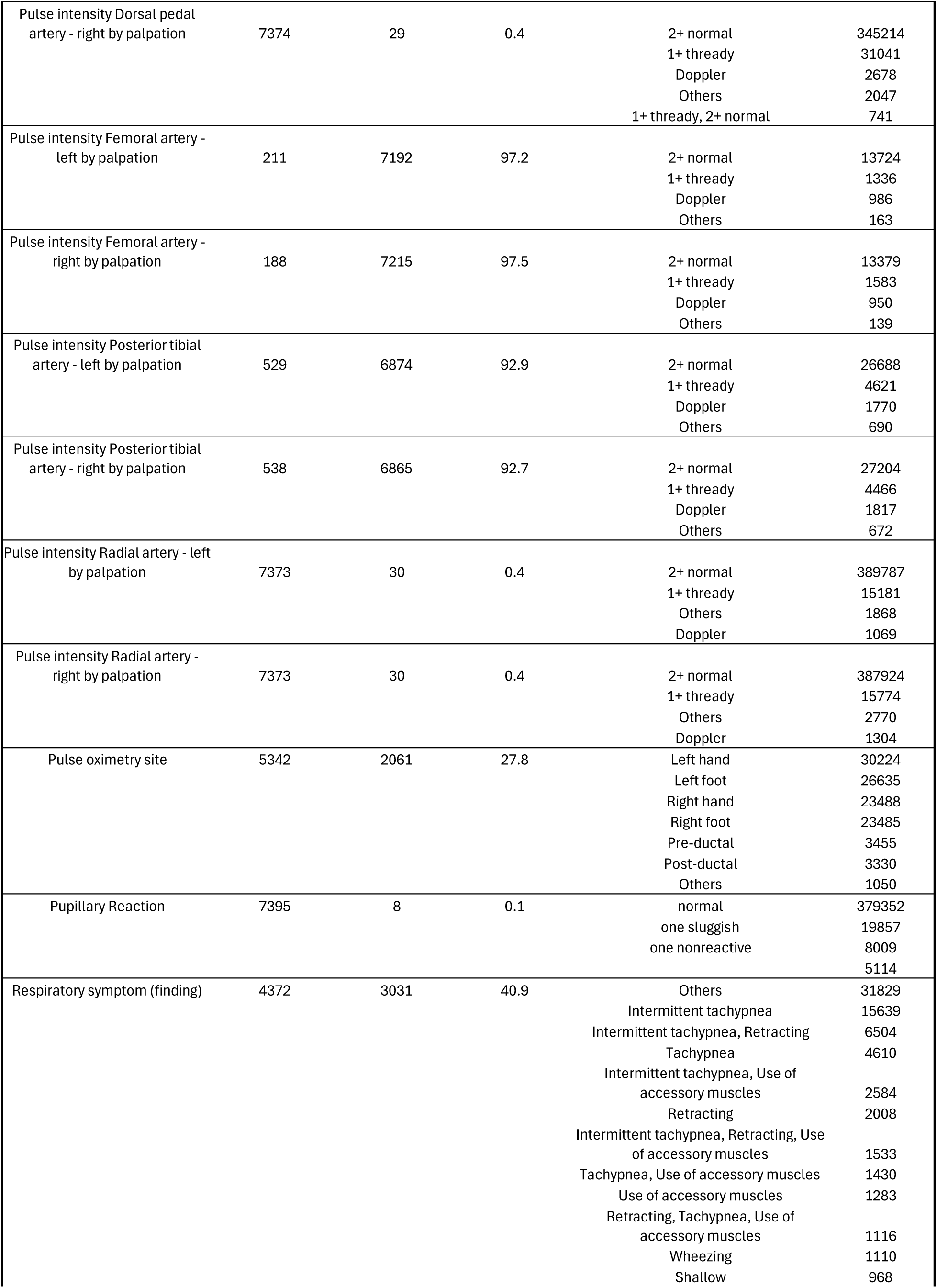
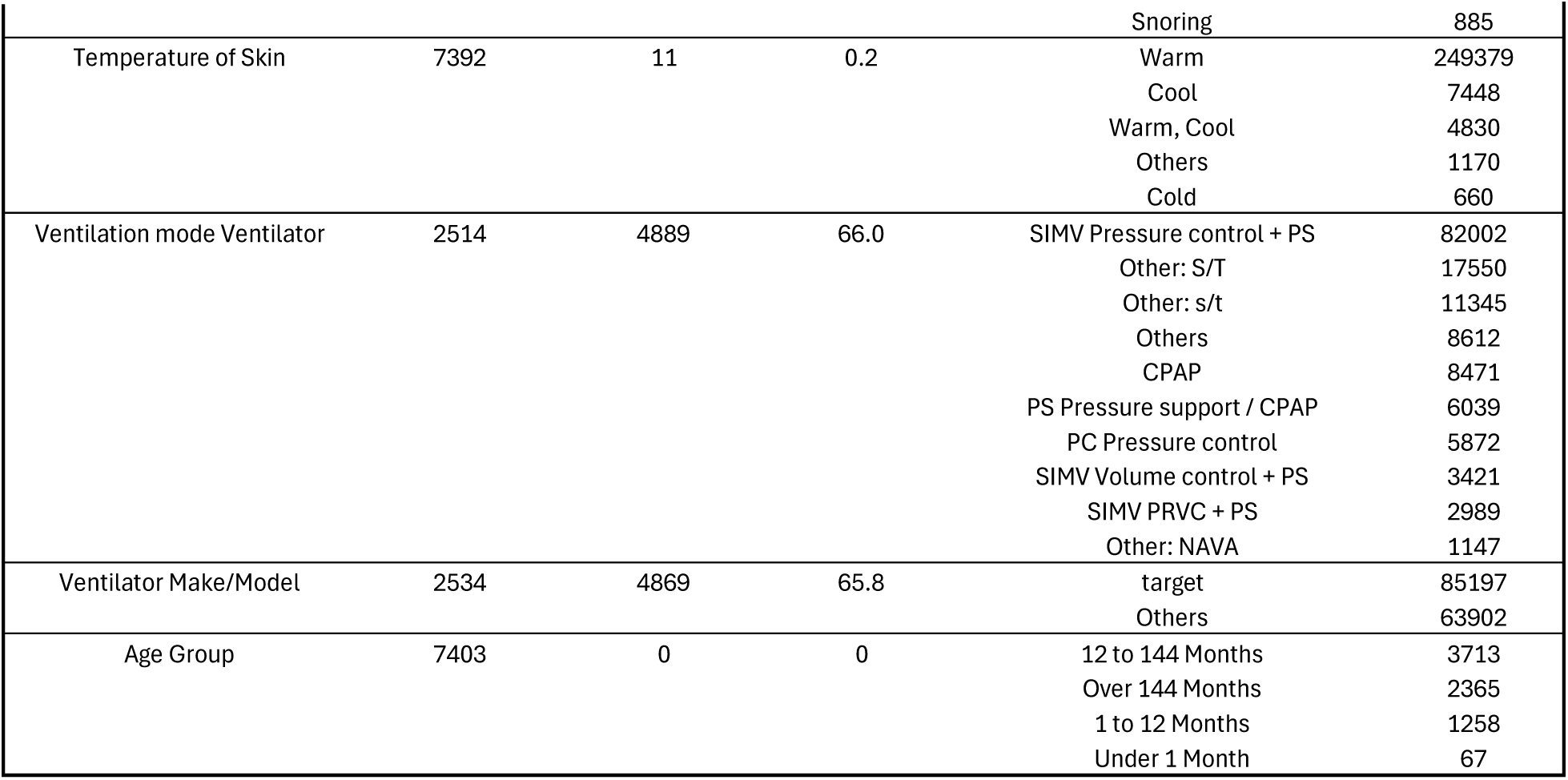
Summary statistics for 53 categorical biomarkers used in the analysis (N = 7403 encounters). For each biomarker, the table reports the number of encounters with at least one recorded value, the number and percentage of encounters missing values, and the recorded category values with their corresponding counts.

**Supplemental Table 5.** Summary statistics for 19 binary biomarkers used in the analysis (N = 7403 encounters). For each biomarker, the table reports the number of encounters with a recorded value (i.e., encounters where the biomarker was ever observed), the number and percentage of encounters missing values (i.e., where the biomarker was not observed or recorded).

| Biomarker_Name | Enc_HasBiomarker<br>(N) | Enc_NoBiomarker<br>(N) | Enc_NoBiomarker<br>(%) |
| --- | --- | --- | --- |
| Care planning and problem solving actions (procedure) | 6700 | 703 | 9.5 |
| Thrombolytic agent not administered because contraindicated (situation) | 202 | 7201 | 97.3 |
| Emotional support management (procedure) | 7333 | 70 | 1.0 |
| Stop time (qualifier value) | 1961 | 5442 | 73.5 |
| Blood product type | 1262 | 6141 | 83.0 |
| Blood product unit ID [#] | 1262 | 6141 | 83.0 |
| Ventilated | 1743 | 5660 | 76.5 |
| Cisatracurium | 571 | 6832 | 92.3 |
| Dobutamine | 10 | 7393 | 99.9 |
| Dopamine | 41 | 7362 | 99.5 |
| Endotracheal tube | 1335 | 6068 | 82.0 |
| Epinephrine | 1159 | 6244 | 84.3 |
| Fentanyl | 1918 | 5485 | 74.1 |
| Hydromorphone | 855 | 6548 | 88.5 |
| Lorazepam | 1757 | 5646 | 76.3 |
| Midazolam | 1965 | 5438 | 73.5 |
| Milrinone | 103 | 7300 | 98.6 |
| Morphine | 1440 | 5963 | 80.6 |
| Norepinephrine | 777 | 6626 | 89.5 |

**Supplemental Table 6.** Summary of biomarker categories and average missingness rates in the encounter level.

| Biomarker Types | Count | Encounter Level<br>Avg Missing(%) |
| --- | --- | --- |
| Lab Values | 78 | 42.3 |
| Nurse Observations / Assessments | 39 | 30.8 |
| Medications | 15 | 87.0 |
| Vital Signs | 14 | 25.0 |
| Device / Therapy Settings | 13 | 47.7 |
| Clinical Procedures / Notes | 5 | 67.9 |
| Demographics | 4 | 39.8 |

Notes:

## Notes

### Lab Values (78 biomarkers)

Alanine aminotransferase [Enzymatic activity/volume] in Serum or Plasma, Albumin [Mass/volume] in Serum or Plasma, Calcium [Mass/volume] in Serum or Plasma, Magnesium [Mass/volume] in Serum or Plasma, Aspartate aminotransferase [Enzymatic activity/volume] in Serum or Plasma, Bilirubin.direct [Mass/volume] in Serum or Plasma, Bilirubin.total [Mass/volume] in Serum or Plasma, Carbon dioxide [Partial pressure] in Exhaled gas --at end expiration, Calcium.ionized [Moles/volume] in Blood, Carbon dioxide [Partial pressure] in Venous blood, Hemoglobin [Mass/volume] in Blood by calculation, Gamma glutamyl transferase [Enzymatic activity/volume] in Serum or Plasma, Osmolality of Serum or Plasma, Oxygen [Partial pressure] in Arterial blood, Oxygen [Partial pressure] in Venous blood, pH of Urine, Phosphate [Mass/volume] in Serum or Plasma, Protein [Mass/volume] in Serum or Plasma, Fibrinogen [Mass/volume] in Platelet poor plasma by Coagulation assay, Platelet mean volume [Entitic volume] in Blood by Automated count, Anion gap in Serum or Plasma, Creatinine renal clearance/1.73 sq M.predicted by Cockcroft-Gault formula, BSA formula, Vancomycin [Mass/volume] in Serum or Plasma –trough, Hematocrit [Volume Fraction] of Blood by Automated count, Specific gravity of Urine by Test strip, Monocytes/100 leukocytes in Blood by Automated count, Prothrombin time (PT) in Blood by Coagulation assay, Alkaline phosphatase [Enzymatic activity/volume] in Serum or Plasma, Beta hydroxybutyrate [Moles/volume] in Serum or Plasma, Basophils [#/volume] in Blood by Automated count, Basophils/100 leukocytes in Blood by Automated count, Eosinophils [#/volume] in Blood by Automated count, Eosinophils/100 leukocytes in Blood by Automated count, Lymphocytes [#/volume] in Blood by Automated count, Variant lymphocytes/100 leukocytes in Blood by Manual count, Lymphocytes/100 leukocytes in Blood by Automated count, Metamyelocytes [#/volume] in Blood by Manual count, Metamyelocytes/100 leukocytes in Blood by Manual count, Monocytes [#/volume] in Blood by Automated count, Myelocytes [#/volume] in Blood by Manual count, Myelocytes/100 leukocytes in Blood by Manual count, Neutrophils [#/volume] in Blood by Automated count, Band form neutrophils [#/volume] in Blood by Manual count, Band form neutrophils/100 leukocytes in Blood by Manual count, Neutrophils/100 leukocytes in Blood by Automated count, MCH [Entitic mass] by Automated count, MCHC [Mass/volume] by Automated count, MCV [Entitic volume] by Automated count, Erythrocyte distribution width [Ratio] by Automated count, Erythrocytes [#/volume] in Blood by Automated count, Blood urea nitrogen, C-Reactive Protein, Chloride, Creatinine, Glucose, Hemoglobin, INR, Lactate, PTT, Platelets, Potassium, Procalcitonin, Sodium, White blood cell count, Pco2, pH, Anisocytosis [Presence] in Blood by Light microscopy, Glucose [Presence] in Urine by Test strip, Hemoglobin [Presence] in Urine by Test strip, Ketones [Presence] in Urine by Test strip, Microcytes [Presence] in Blood by Light microscopy, Polychromasia [Presence] in Blood by Light microscopy, Protein [Presence] in Urine by Test strip, Differential cell count method – Blood, Base deficit, Base excess, Bicarbonate, ABO and Rh group [Type] in Blood from Donor.

### Nursing Observations / Assessments (39 biomarkers)

Humpty Dumpty fall risk assessment score (observable entity), Body position (observable entity), Capillary refill [Time] of Nail bed, Cardiac rhythm NEMSIS, Heart rate rhythm, Character of cough (observable entity), Character of pulse (observable entity), Color of Stool, Volume of Stool, Color of skin (observable entity), Diet followed (observable entity), Gait type (observable entity), Glasgow coma score eye opening, Glasgow coma score motor, Glasgow coma score verbal, Level of consciousness, Muscle tone (observable entity), Mouth care (regime/therapy), Pain severity total [Score] FLACC, Pain severity Activity FLACC, Pain severity Cry FLACC, Pain severity Face FLACC, Pain severity Wong-Baker FACES pain rating scale, Pulse intensity Brachial artery - left by palpation, Pulse intensity Brachial artery - right by palpation, Pulse intensity Dorsal pedal artery - left by palpation, Pulse intensity Dorsal pedal artery - right by palpation, Pulse intensity Femoral artery - left by palpation, Pulse intensity Femoral artery - right by palpation, Pulse intensity Posterior tibial artery - left by palpation, Pulse intensity Posterior tibial artery - right by palpation, Pulse intensity Radial artery - left by palpation, Pulse intensity Radial artery - right by palpation, Pupillary Reaction, Respiratory symptom (finding), Peds Coma Score, Ability to perform mouthcare activities (observable entity), Affect function (observable entity), Passing flatus (finding).

### Medications (15 biomarkers)

Tacrolimus [Mass/volume] in Blood by LC/MS/MS, Cisatracurium, Dobutamine, Dopamine, Epinephrine, Fentanyl, Hydromorphone, Lorazepam, Midazolam, Milrinone, Morphine, Norepinephrine, Blood product type, Blood product unit ID [#], Packed erythrocytes given [Volume].

### Vital Signs (14 biomarkers)

Temperature, Pulse, Respiratory Rate, SBP, DBP, SpO₂, Temperature of Skin, Body weight --used for drug calculation, Body mass index (BMI) [Ratio], Intracranial pressure (ICP), Central venous pressure (CVP), Cerebral perfusion pressure, Weight, MBP.

### Device / Therapy Settings (13 biomarkers)

Positive end expiratory pressure setting Ventilator, Tidal volume setting Ventilator, Tidal volume expired Respiratory system airway --on ventilator, Ventilation mode Ventilator, Ventilator Make/Model, Pulse oximetry site, Blood pressure measurement site, Heart rate measurement site, Ventilated, Endotracheal tube, Body temperature measurement site, Inhaled oxygen concentration, Inhaled oxygen flow rate.

### Clinical Procedures / Notes (5 biomarkers)

History and physical note, Care planning and problem solving actions (procedure), Thrombolytic agent not administered because contraindicated (situation), Emotional support management (procedure), Stop time (qualifier value).

### Demographics (4 biomarkers)

Gestational age--at birth, Body surface area, Body height, Age Group.

